# Air pollutant multiomics improves functional annotation of SNPs associated with lung disease

**DOI:** 10.64898/2026.06.21.26356065

**Authors:** Hope A. Townsend, Sarah K. Sasse, Shu Yi Liao, Anthony N. Gerber, Robin D. Dowell, Arnav Gupta

## Abstract

Particulate matter exposure impacts asthma and chronic obstructive pulmonary disease (COPD) outcomes, and this risk is expected to grow. Therapies that target disease and exposure risk mechanisms are necessary to address this concern. We identified airway epithelial transcriptional mechanisms associated with exposure and disease risk using a functional genomics pipeline that combined unbiased nascent RNA sequencing, genetic prioritization, and gene-regulatory element correlation. We then applied functional studies to prioritized regulatory elements and transcriptional mechanisms and established hyaluronic acid metabolism and *TOMM7* regulation as plausible targets supported by airway models and primary human data. Our pipeline shifts the objectives of exposure and genetic studies from broad discovery to refining specific targets, which are more focused on therapeutic manipulation. While future studies are necessary to validate the targets, our framework integrates exposures, transcriptional biology, and human genetics to improve prioritization and streamline translation between target discovery and validation.

**Graphical Abstract:** 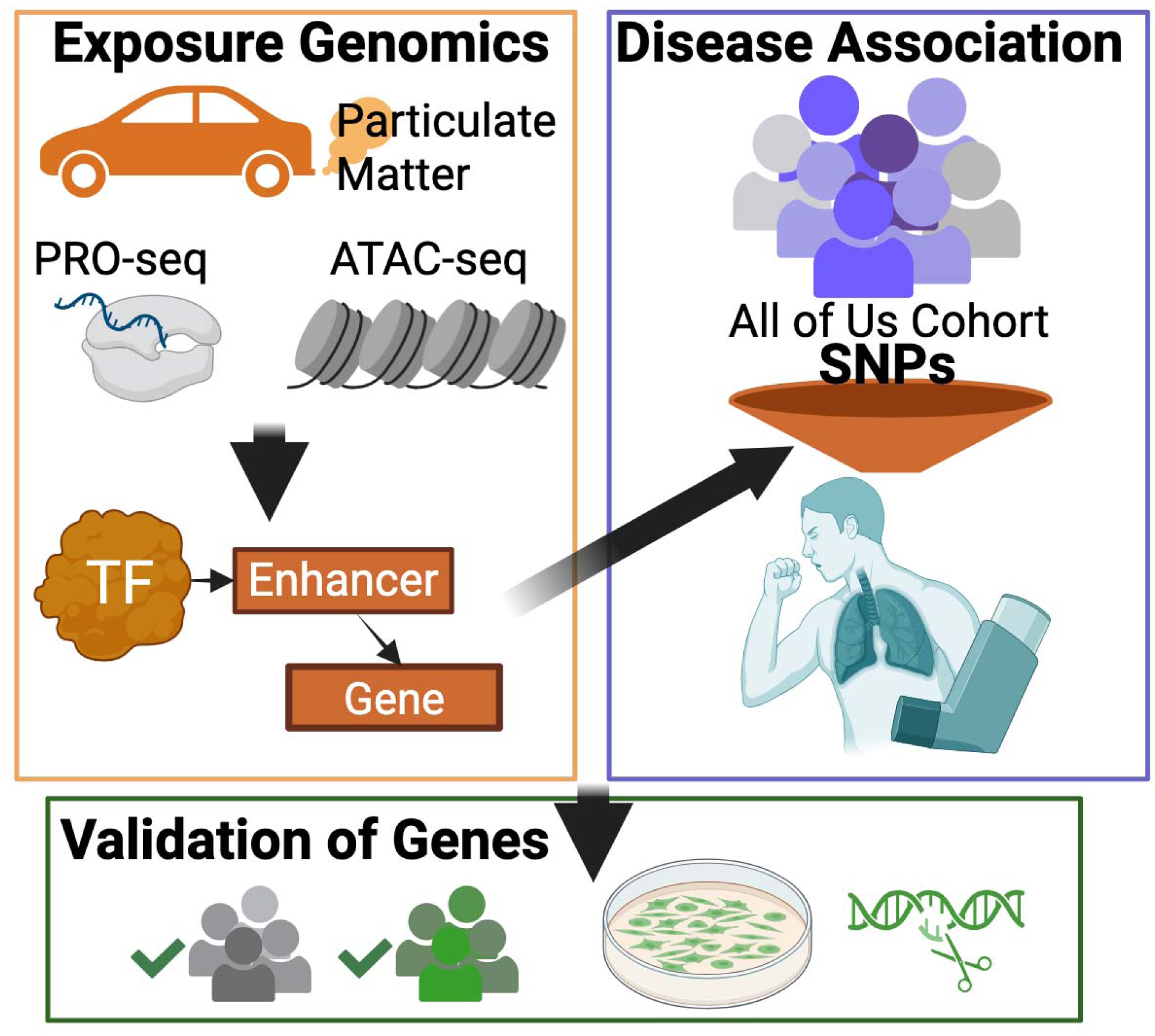

## Introduction

Asthma and chronic obstructive pulmonary disease (COPD) are complex diseases affecting approximately 15% of adults in the United States (*Centers for Disease Control and Prevention*, 2023). Adverse asthma and COPD outcomes result directly from inhaled exposures, such as particulate air pollution (Sin et al., 2023). Environmental particulate matter (PM) is expected to increase in magnitude over the next 30 years due to wildfire smoke and climate change (Foundation, 2022; Proestakis et al., 2025). Addressing this growing health concern requires understanding how individuals with disease respond to exposure and who are at the greatest risk of adverse outcomes. However, exposure response mechanisms and individual risk are poorly understood, and precision medicine strategies to prevent the impact of particulate exposures on asthma and COPD remain limited. Detailed studies that connect specific genes, individual risk factors, and cellular mechanisms of disease are necessary to address these knowledge gaps.

Prior exposure studies lack the resolution to define detailed disease mechanisms, which may include rapidly regulated transcriptional responses. Nascent RNA sequencing using precision run-on sequencing (PRO-seq) enables quantification of rapid responses, which are enriched in disease-relevant mechanisms, such as aryl hydrocarbon receptor (AHR) signaling (Gupta et al., 2021). Even minor perturbations in these rapid responses may affect disease risk since chronic diseases, such as asthma and COPD, result from repeated exposures. Additionally, PRO-seq captures transcriptional activity in non-coding regulatory regions, which can be used to infer transcription factor activity or connect regulatory regions with gene targets (Rubin et al., 2021; Sigauke et al., 2025; H. A. Townsend et al., 2026). Therefore, we hypothesize that nascent transcriptional analysis will reveal novel mechanistic details linking exposures to disease.

Genetic study is another tool that can propose mechanistic details based on the association between genome sequence variants and disease risk in the population. However, despite the high heritability of COPD and asthma, genome-wide association studies (GWASs) have yielded limited mechanistic understanding, particularly in the context of environmental exposures (Cho et al., 2022a; J. J. Zhou et al., 2013). Several limitations are responsible for this knowledge gap. First, GWASs that include exposures are vastly underpowered to detect exposure-genetic interactions (Cho et al., 2022b), and the timing and dose dependence of exposures on gene expression responses likely contributes to this limitation. Second, statistical prioritization in most GWASs has led to high false-negative rates (Schaid et al., 2018). Indeed, non-coding single-nucleotide polymorphisms (SNPs), which represent 90% of disease risk (Chen et al., 2020), often have low individual effect sizes (Stringer et al., 2011). Third, GWASs typically limit analysis to the SNP with the highest statistical association, missing critical functional SNPs that define disease biology (Schaid et al., 2018). Fine-mapping SNPs with multiomics data has improved the ability to prioritize function over statistics and link SNPs to biological mechanisms (Cho et al., 2022a; Gu et al., 2023; Sasse et al., 2025), but multiomics tools rarely address non-coding SNPs and are limited to general databases rather than disease-specific experiments (Moore et al., 2020; Watanabe et al., 2017). In contrast, combining exposure multiomic experiments, especially methods like PRO-seq that can annotate non-coding regions, with genetic association links transcriptional networks, non-coding SNPs, and genes that define detailed mechanisms of disease.

We applied a functional genomics pipeline that leverages nascent RNA sequencing and genetics to identify specific genes and mechanisms that are relevant to exposures and disease. We first compared nascent RNA responses in airway epithelial systems at short exposure durations (30 and 120 minutes) between two PM models – wood smoke particles (WSP) (Gupta et al., 2021) and urban particulate matter (UPM). In addition to conventional bulk analysis using gene ontology and motif enrichment, we applied a focused analysis that used genetic variants and multiomics annotations to prioritize regulatory elements for functional analysis. We used published genomic data sets and quantitative trait locus analyses to further select candidates based on genomic functions. Finally, we bioinformatically predicted genes regulated by high-priority regulatory elements using nascent RNA data and tested one prediction using CRISPR genome editing. Our work defined a precise set of SNPs, regulatory elements, and genes that contribute important details to mechanistic links between PM exposures, asthma, and COPD.

## Results

### Nascent transcript sequencing with PRO-seq identified direct transcriptional targets of UPM

Airway epithelial cells exposed to PM exhibit temporally dynamic responses (Giammona et al., 2025; Gupta et al., 2021, 2022; N. Kim et al., 2019). We first applied PRO-seq at two early time points (30 and 120 minutes) following UPM exposure in primary small airway epithelial cells (smAECs). Early time points were selected to minimize trans-regulatory effects and capture rapid-transient responses that we previously observed in exposure PRO-seq data at identical time points (Gupta et al., 2021). Transcriptional responses were separated into three groups based on temporal response within the measured time points: early (significant change at 30 minutes, return to baseline at 120 minutes), early-plateau (significant change at 30 minutes, sustained at 120 minutes), and late (significant change only after 120 minutes) (Zenodo). The largest number of differentially transcribed genes were found in the early response category, and based on gene ontology annotations, cellular pathways involved in the direct UPM response included cell adhesion and hyaluronic acid biosynthesis (Figure 1A-B, Supplemental Figure 1).

**Figure 1.**
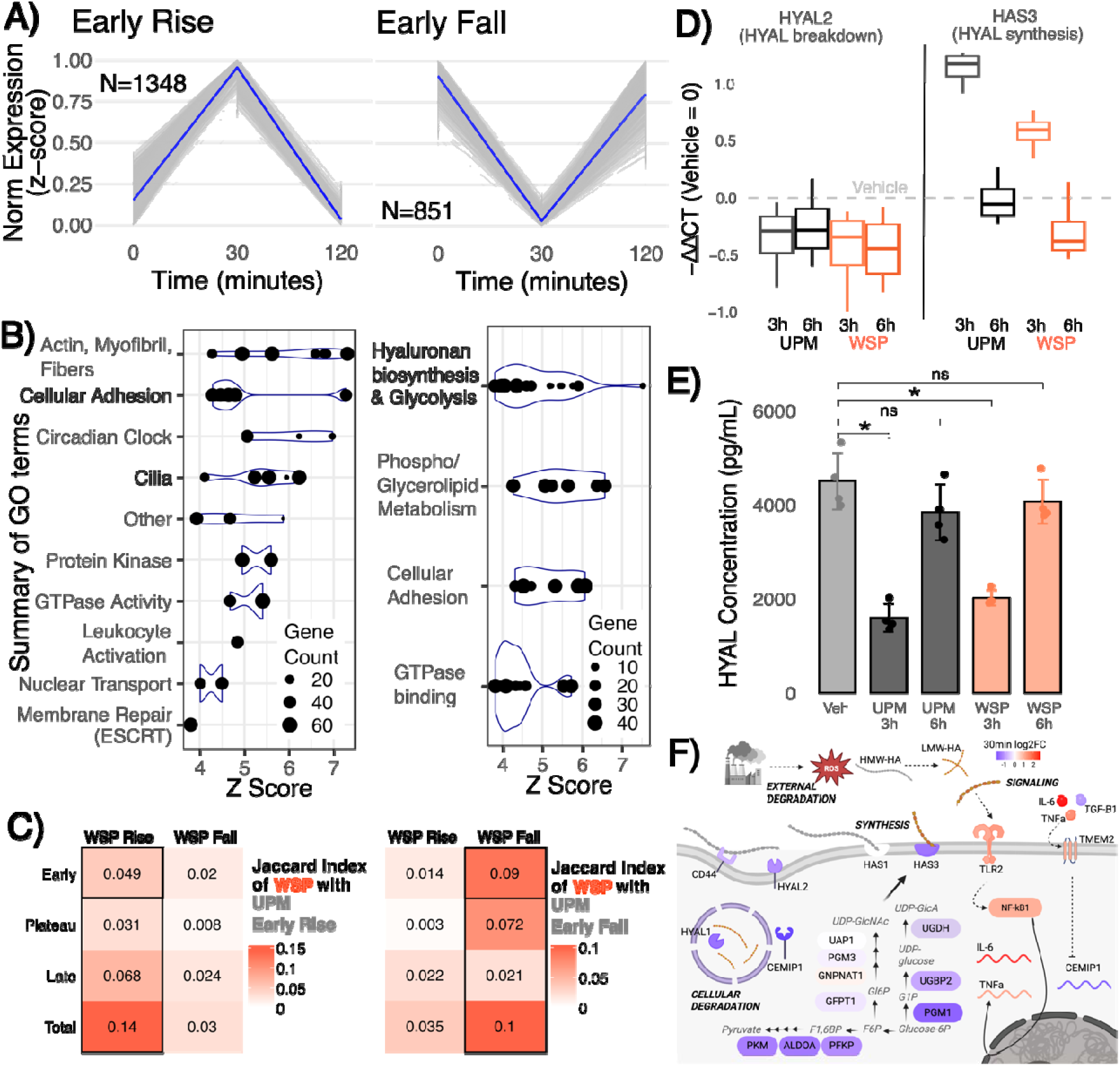
UPM produces a timeline of responses comparable to other particulate perturbations. **A)** Line plots of the mean (blue, light blue 95% confidence interval) and individual (grey) normalized transcriptional changes of statistically significant genes in early response. Norm Expression (z-score) =min-max scaled normalized counts. 0 minutes is Vehicle. **B)** Summary of significant Gene Ontology (GO) terms of the matching genes with dot size indicating the number of significant genes matching the GO term. **C).** Jaccard index comparing UPM early-response genes with WSP transcriptional responses. **D)** qRT-PCR or **E)** ELISA (hyaluronic acid levels in media) results from smAECs perturbed with UPM or WSP for 3 and 6 hours. **F)** Log_2_ Fold Change expression at UPM 30 min for genes involved in hyaluronic acid degradation, synthesis, and related signaling. H/LMW-HA=high/low molecular weight hyaluronic acid. ROS=reactive oxygen species. Adjusted p-value < 1x10^-10^ was used.

We then compared UPM with WSP exposure in Beas-2B airway epithelial cells (Gupta et al., 2021) at identical 30- and 120-minute durations to determine if we could separate unique and shared transcriptional responses between exposures in a related cell system, as we have previously observed similar exposure responses between Beas-2B and smAEC models (Gupta et al., 2021, 2022). There was significant overlap in the differentially transcribed genes between UPM and WSP exposures while the temporal dynamics were less aligned (Figure 1C). These findings demonstrated that PRO-seq could separate specific transcriptional responses that were shared or distinct between multiple PM sources.

### Nascent transcriptional responses are associated with mRNA and functional changes in airway epithelial cells exposed to PM

To determine if genes in the early-response time category, which contained the largest number of responses but returned to baseline within 120 minutes, had sustained biological relevance, we next examined whether these genes were also regulated at the mRNA and functional levels. We first re-analyzed published RNA-seq data for WSP and found that genes transcribed at the 30-minute PRO-seq time points had the greatest correlation with 2-hour RNA-seq data based on log_2_ fold change (Spearman rho = 0.39; Supplemental Figure Corr_Comp A). For detailed functional validation, we focused on hyaluronic acid metabolism. In addition to strong enrichment in gene ontology within the early response category (GO:0030213 hyaluronan biosynthetic process, q = 0.008, fold change = 16.8, fold enrichment rank in early fall UPM category = 1), hyaluronic acid metabolism has prior connections with inhaled exposures, asthma and COPD risk, and potential therapeutic manipulation (Galdi et al., 2021; Papakonstantinou et al., 2014; Petrigni & Allegra, 2006; Waeijen-Smit et al., 2021). We exposed smAECs from 2 donors to UPM and WSP and measured gene expression by qRT-PCR and functional signal by Enzyme Linked Immunosorbent Assay (ELISA) for regulated targets at time points that were previously established to correlate with nascent RNA responses. In nascent transcription, genes involved in hyaluronic acid breakdown (*HYAL1, HYAL2, HYAL3*) had reduced transcriptional signal at 30 minutes of UPM or WSP exposure (Supplemental Figure HYA), and we also observed reduced mRNA expression of *HYAL1*, *HYAL2*, and *HYAL3* by qRT-PCR (Figure 1D, Supplemental Figure HYA_qRTPCR). Nascent transcription and steady-state mRNA levels were less correlated for hyaluronic acid synthesis genes (*HAS2*, *HAS3*). While UPM decreased *HAS3* nascent transcription after 30 minutes and WSP increased *HAS3* nascent transcription after 30 minutes, *HAS3* steady-state mRNA expression was increased by both exposures at 3 hours (Figure 1D, Supplemental Figure HYA). *HAS2* also showed increased steady-state mRNA expression by qRT-PCR following both exposures (Supplemental Figure HYA_qRTPCR). Comparisons between nascent and mRNA responses supported the roles for hyaluronic acid metabolism and other biologic processes identified by transient nascent RNA to identify cellular mechanisms associated with PM exposures.

We next tested whether transient nascent RNA responses had implications in cellular or disease contexts, again using hyaluronic acid metabolism as a representative example due availability in cell models and human plasma. First, we observed an acute decrease in total hyaluronic acid in the culture media of smAECs exposed to UPM or WSP at 3 hours followed by a return to baseline by 6 hours of exposure (Figure 1E). Then, we found that a 1 unit increase in hyaluronidase-1 plasma protein levels predicted increased Forced Expiratory Volume in 1 second (FEV1) change (ml/yr) by 0.0024 (high-precision p-value 0.0016) in a linear regression model of the Genetic Epidemiology of COPD cohort (COPDGene, N=4,857 individuals) after adjusting for smoking status, gender, age, BMI, and total cumulative cigarette exposure. These findings demonstrated that nascent transcription, even in rapid and transient responses, identified genes that are relevant to cell physiology and disease biology.

Finally, we also compared nascent and mRNA responses within inflammatory genes, which are established responses to PM exposures (Gupta et al., 2021) and interact with both hyaluronic acid metabolism and COPD risk (Campo et al., 2012; Sato et al., 2021, 2023; Waeijen-Smit et al., 2021). We examined transcriptional responses of genes encoding toll like receptors (TLR2 and TLR4), cytokines (IL-6, TNF-alpha, and TGF-beta), and additional regulators of hyaluronic acid (*CEMIP1* and *TMEM*) and observed consistent agreement between nascent transcription and mRNA expression (Figure 1F, Supplemental Figure HYA). While these findings strengthened the relevance of transient nascent RNA responses to PM exposures and regulation of hyaluronic acid, they also motivated additional study of transcriptional networks to determine upstream regulators of PM responses.

### Network analysis identifies distinct transcriptional programs that regulate particulate matter exposures

To further understand the relevance of early-response transcription in exposure and disease biology, we applied Transcription Factor Enrichment Analysis (TFEA) (Rubin et al., 2021; H. A. Townsend et al., 2026) to transcribed regulatory elements (tREs, also known as enhancers or cis-regulatory elements) and defined transcription factor regulators of the early transcriptional response. Similar to gene transcription, we separated tRE responses to UPM into early, early-plateau, and late temporal categories and observed agreement in tRE transcription between UPM and WSP exposures (Supplemental Figure EnhancerResponse). To determine if the transcription factors directly regulating the early transcriptional response were relevant to exposure and disease biology, we applied TFEA in samples exposed to UPM or WSP for 30 minutes (Figure 2A, Supplemental Tables 11-12). The presence of several transcription factors previously implicated in exposure and disease biology, such as AHR/AHRR, NFKB, and EGR (Beamer & Shepherd, 2013; Cui et al., 2025; Zou & Zeng, 2023), validated the importance of early-response transcriptional regulation. In comparison, at the 120-minute exposure time point, these relevant transcription factors were not significantly enriched and fewer transcription factors were identified by TFEA overall (Supplemental Figure TFEAscatter B).

**Figure 2.**
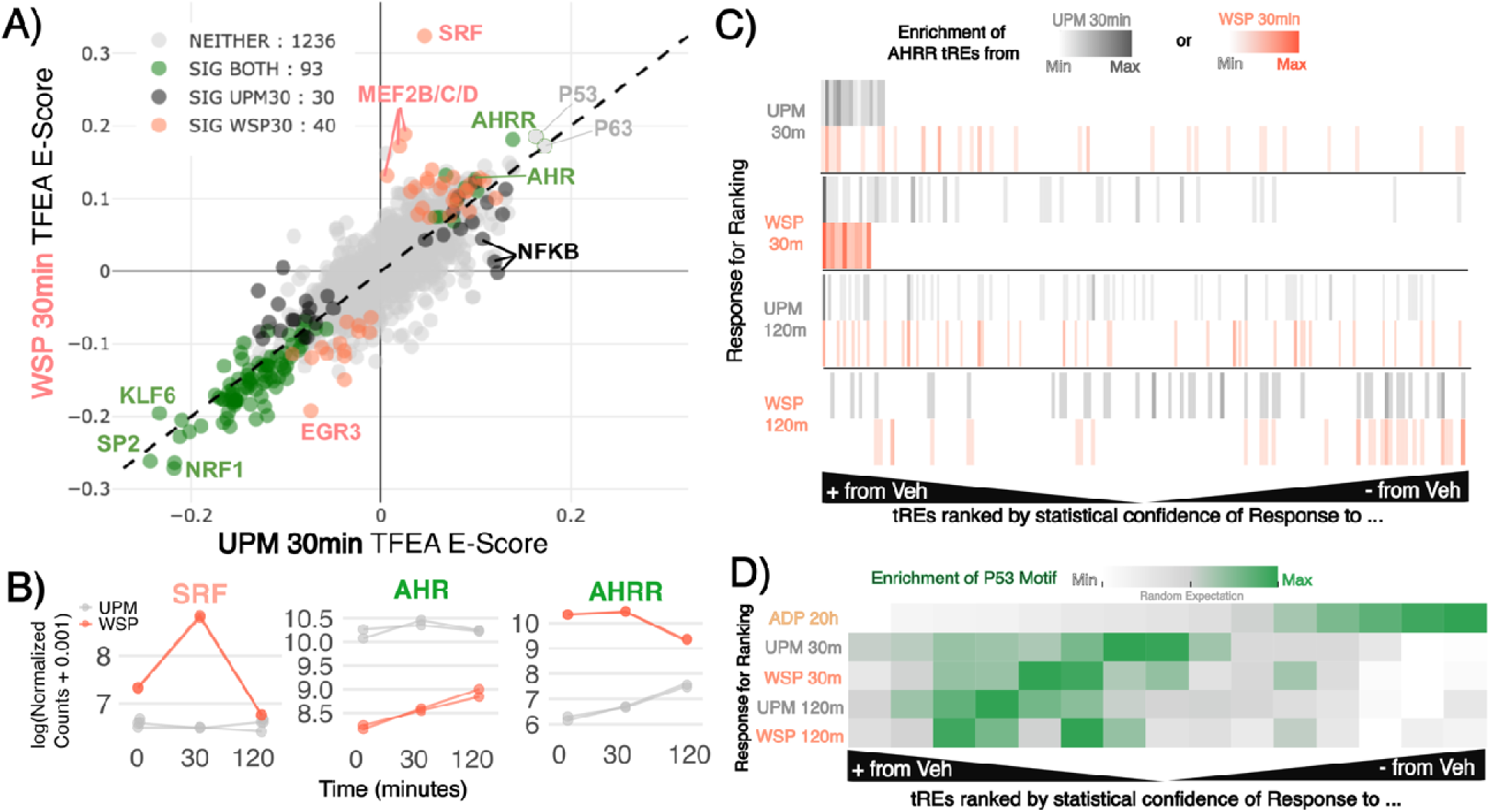
UPM produces a specific response of transcriptional networks (transcription factors and corresponding tREs). **A)** Scatterplot of TFEA GC-corrected enrichment scores for WSP and UPM 30 minutes. Transcription factors are highlighted as being a significant call in neither UPM/WSP, only one, or both (see Methods) **B)** Log normalized counts of transcription factor genes in UPM and WSP treated cells across time. **C)** tREs called responding via AhRR to WSP and UPM at 30 minutes are colored pink and grey, respectively. The x-axes are rankings of tREs according to DESeq2 p-value (poles=lowest) of up(+) or down(-) regulation compared to Vehicle across 4 different responses: WSP 30min, WSP 120min, UPM 30min, UPM 120min. **D).** Quantiles of tREs are ranked as done in C for 5 different responses: ADP 20hr, WSP 120min, UPM 30min, UPM 120min. Relative enrichment of the P53 motif (weighted score from TFEA) is colored within each quantile (quantile N=2,598, scaled within each row) where green shows maximum enrichment.

We next examined the potential for early response transcription factors to influence disease risk through positive feedback regulation and chromatin priming. To evaluate the potential for positive feedback, we compared the transcriptional kinetics of the gene encoding the transcription factor with its motif enrichment. Transcription factors independent to each exposure, such as SRF for WSP and RELB or NFKB2 for UPM, showed increases in both gene signal and tRE motif enrichment, suggesting a potential for positive feedback regulation (Figure 2B, Supplemental Figure TFlevels). Next, since changes in chromatin accessibility can contribute to disease risk by priming cells for future exposures, we applied TFEA and the assay for transposase accessible chromatin with sequencing (ATAC-seq) in cells exposed to UPM and WSP for 30 minutes (Supplemental Figure TFEAscatter A, Supplemental Tables 13-14). We observed limited correlation in log_2_ fold changes of tRE transcription (PRO-seq) and accessibility (ATAC-seq) – maximum Spearman rho = 0.35 – compared to correlation across perturbations within the same assay (maximum Spearman rho = 0.64 between ATAC-seq 30 min) (Supplemental Figure Corr B). Yet TF motif enrichment scores from TFEA were still highly similar between PRO-seq and ATAC-seq at the 30min mark (Spearman rhos > 0.8; Supplemental Figure Corr C). Specifically, multiple transcription factors, such as AHR/AHRR and KLF, showed enriched motifs in tREs changing transcription (PRO-seq) or accessibility (ATAC-seq) and were previously associated with exposures and disease (G.-D. Kim et al., 2024; Wan et al., 2024), supporting the relevance of early changes in chromatin access. AHR and AHRR putatively regulated both early transcriptional and chromatin accessibility responses to UPM and WSP exposures, but we observed subtle differences in *AHR* and *AHRR* gene transcription, suggesting differences AHR/AHRR activity based on exposure or cellular context (Figure 2B). We explored these differences further by identifying tREs putatively regulated by AHRR based on increased nascent transcription after 30 minutes of UPM or WSP exposure using the TFEA-leading edge method (H. A. Townsend et al., 2026). UPM-responsive AHRR tREs had minimal overlap with WSP-responsive AHRR tREs, suggesting separate regulation of AHRR-driven nascent transcriptional responses between UPM and WSP (Figure 2C). For comparison, the early-rise pattern observed for WSP-specific AHRR tREs was similar to that of tREs attributed to WSP-specific transcription factor SRF (Supplemental Figure SRFtREs). No major differences in nascent transcription of canonical AHR/AHRR target genes (Gupta et al., 2021) were observed between UPM and WSP exposures (Supplemental Figure AhRTargets).

Finally, to evaluate the long-term relevance of transcription factors that regulated rapid responses, we compared TFEA results between early (30 or 120 minutes) and late (20 hours) time points. We selected P53 to compare motif enrichment between UPM, WSP, and Afghan Desert Particulates (ADP) exposures because P53 has been previously reported as a transcription factor shared across multiple PM models and associated with disease (Gupta et al., 2022; Mizuno et al., 2017; Sagiv et al., 2018, p. 53; Soberanes et al., 2006; W. Zhou et al., 2016). We observed P53 enrichment in tREs with a weakly increased signal at both 30 and 120 minutes of WSP and UPM exposure and a strongly reduced signal at 20 hours of ADP exposure (Figure 2D). While the direction of association between P53 enrichment and tRE responses changes, these data suggest that regulators of rapid responses are also enriched at later time points, further supporting the role of rapid responses in affecting cellular biology. However, our results also showed variations in tRE regulation by the same transcription factor. Therefore, focused analysis of tREs was necessary to determine the precise transcriptional mechanisms that link exposures with disease.

### SNPs associated with COPD and asthma highlight specific PM responses involved in disease

We used population cohort data to identify which PM exposure responses in airway epithelial cells could be genetically associated with COPD and asthma risk. We specifically asked whether SNPs within PM-responsive tREs were associated with COPD or asthma and linked these SNPs back to the impacted tREs, transcriptional networks, and genes using multiomics data (Figure 3A). We captured full RNA coordinates for 15,136 tREs that responded to PM exposures (Supplemental Figure UpSetResponse) and evaluated a total of 20,764,608 bp (28% +/-1 kb transcription start site (TSS), 36% intragenic, 36% intergenic) of the regulatory genome. The All of Us cohort (The All of Us Research Program Investigators, 2019) had 6,164 SNPs with minor allele frequency ≥0.05, ≥90% completion rate, and mapping within the ∼20 Mb of evaluated regulatory regions. Of the 6,164 SNPs, none had a Benjamini-Hochberg False Discovery Rate (FDR) q value of < 0.05 but 681 had nominal associations (p < 0.05) with COPD or asthma (Figure 3B). SNPs, functional annotations, and additional details (linkage disequilibrium clumping, minor allele frequencies, and p and q values) are listed in Supplemental Tables 3, 4, and 5.

**Figure 3.**
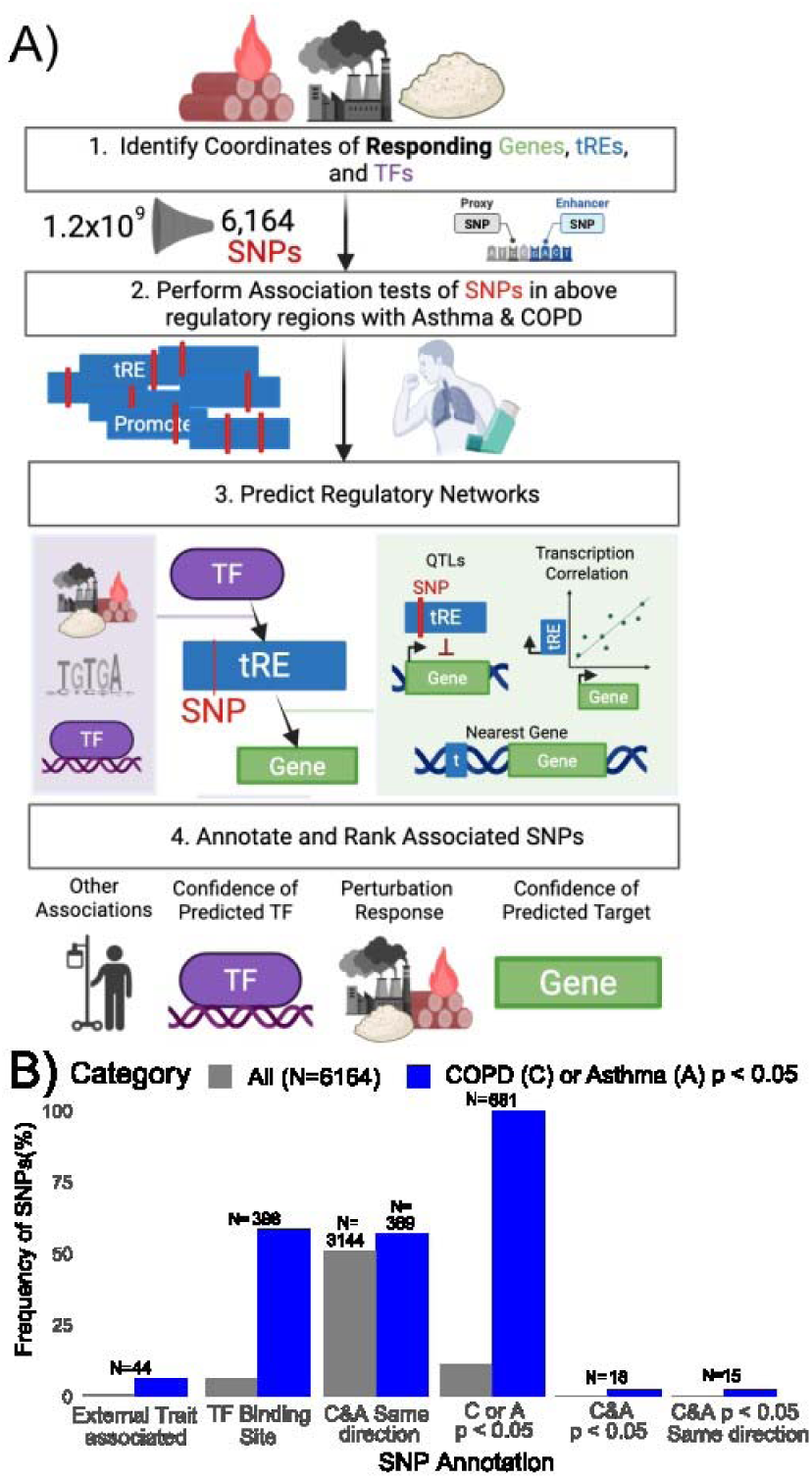
Perturbation-focused regulatory region SNPs associated with COPD and asthma are externally associated with other lung-related traits. **A)** Visualization of pipeline. 1) Identify the genes, tREs, and transcription factors responding to air pollutant perturbations and the coordinates of TSSs and tREs. 2) Evaluate the association of SNPs within these coordinates and with asthma and COPD in All of Us. 3) Predict transcription factor-tRE linkages by considering the transcription factors responding to perturbations, motifs within SNPs, and ChIP-seq binding sites. Predict gene-tRE linkages based on the nearest gene, quantitative trait loci, and correlation of transcription across lung cells. 4) Rank candidate SNPs based on associations, regulatory networks, and tRE response to perturbations. **B)** Percentage of all region SNPs (grey) vs those associated with COPD (C) or asthma (A) (blue) that fall into different functional annotations (details in Methods).

To derive the biologic pathways impacted by these SNPs, we leveraged regulatory network analysis of nascent transcription and chromatin accessibility responses to PM exposures. These networks predicted upstream transcription factors and downstream gene targets of tREs containing SNPs. In addition to the exposure multiomics data, we integrated external genomic data from ENCODE (Moore et al., 2020) and nascent RNA sequencing data sets of airway epithelial cells (Sigauke et al., 2025). We then prioritized SNPs with the strongest support for downstream validation based on association with COPD or asthma, previously found association with traits in ClinVar or GWAS Catalog, confidence of regulatory network support, and response to particulate matter (Figure 3A Step 4; details in Methods) (Cerezo et al., 2025; Landrum et al., 2018). These regulatory network annotations provided biologic validation for associated SNPs and guided the interpretation of specific SNPs prioritized in subsequent experiments.

Next, we used summary statistics from a GWAS of lung function measurements (Shrine et al., 2023a) to evaluate the generalizability of the 681 prioritized variants. After pruning variants for linkage disequilibrium and harmonizing with variants coded in the summary statistics, we compared the remaining 600 SNPs from our results in aggregate against randomly permuted blocks of 600 SNPs matched on minor allele frequency. We compared the mean -1og_10_ P value for association with two lung function measurements – Forced Expiratory Volume in 1 second (FEV1) and FEV1/Forced Vital Capacity (FVC) – between the candidate set and all permuted sets and observed that the candidate set had a greater average statistical value (FEV1 null confidence interval = 0.73 – 1.03, observed = 1.22, q = 2.5 x 10^-4^; FEV1/FVC null confidence interval = 0.68 – 1.02, observed = 1.31, q = 4.8 x 10^-4^). Next, we evaluated SNPs individually using external summary statistics from this GWAS. Using a FDR q < 0.05 threshold within the set of 681 variants, 56 SNPs had significant associations for FEV1 or FEV1/FVC (Supplemental Table 15). This replication analysis provided statistical support for applying the 681 variants as a filter to select tREs for further analysis. However, before further downstream experiments, we evaluated the relevance of rapid-transient transcriptional responses in the context of these genetic signals.

### Rapid-transient nascent transcriptional responses were enriched with disease SNPs and correlated with steady-state RNA responses

To determine if rapid-transient transcriptional responses contained relevant genetic signals, we compared SNP enrichment and correlation with steady-state RNA-sequencing across time categories. Whether considering transcription (PRO-seq) or chromatin accessibility (ATAC-seq), we observed comparable enrichment of SNPs associated with COPD or asthma across all time categories (Supplemental Figure SNP_sig). We then compared steady-state RNA changes from RNA-seq (2h and 4h) (Gupta et al., 2025) to the nascent transcription changes from PRO-seq (30m and 120m) in Beas-2B cells perturbed with WSP. These time points were again selected based on established correlations between nascent and mRNA transcription within this cell type (Gupta et al., 2021). Regardless of time category, about 50% of nascent-responding genes also changed in steady-state levels, with transient genes showing the highest percentage changing in the same direction at steady-state (Supplemental Figure RNAseq_Comp). These findings demonstrated the relevance of genetic signals found in rapid transient transcriptional programs.

### Enhancer-focused analysis compares SNP significance across cohorts and within linked regions

Next, we performed targeted multiomics analysis in genomic regions with replicated genetic signals. First, we leveraged multiomics data to connect the SNPs that we associated with disease in All of Us to additional SNPs based on shared tRE functions (Figure 4A). We demonstrated one example of this approach at a 30 kb locus on chromosome 6, where we identified 2 SNPs in early response tREs (rs3095340 and rs9468829) associated with asthma in All of Us (OR = 1.03, p = 0.02 and 0.04, respectively). The rs3095340 also had significant associations with FEV1 (q = 6.64 x 10^-19^) and FEV1/FVC (q = 2.06 x 10^-13^) in lung function summary statistics. Another early response tRE at this locus contained rs3094117, which was significantly associated with asthma in the Genetic Epidemiology Research on Aging (GERA) cohort (Sasse et al., 2025) (Figure 4B). All three tREs shared PRO-seq and ATAC-seq signal trends across time-points, and ChIP-seq peaks for GR, JUN, GABPA, and FOSL2 in pulmonary epithelial models, representing potential transcriptional regulators underlying the disease risk (Supplemental Figure DEX SNP). We found a similar case of four SNPs within three overlapping tREs in a 5 kb locus on chromosome 12. In this case, these four SNPs were ranked within the top 10 genetic associations with COPD or asthma in our study (Supplemental Figure Chr12SNPs). The relevance of these SNPs was supported by population allele frequencies across multiple ancestries suggesting a possible fitness advantage associated with the alternate allele, following the same direction as the odds ratios for COPD and asthma risk for these SNPs in All of Us (Supplemental Figure Ppln). Therefore, multiomic analysis of tRE clusters improved replication across multiple cohorts and multiple ancestries and suggested underlying transcriptional regulatory mechanisms of disease.

**Figure 4.**
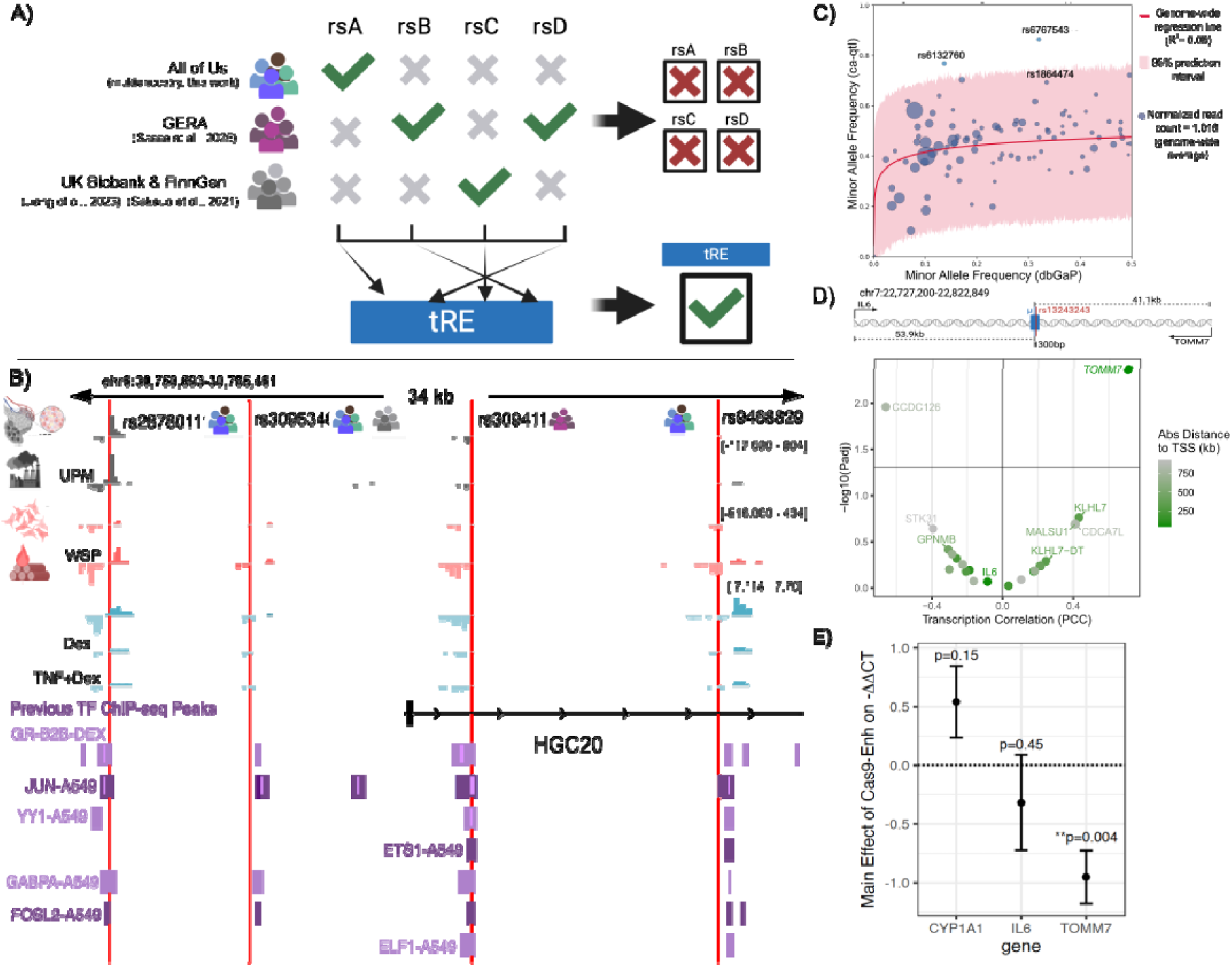
Multiomic and functional analysis of high priority SNPs. **A)** Individual SNPs may not be replicated across cohorts while the tRE to which SNPs link does. **B)** IGV tracks for SNP-enriched tRE groups: PRO -seq of smAEC perturbed with UPM and Beas-2B cells perturbed with WSP, and GRO-seq of Beas-2B cells perturbed with dexamethasone (Dex), TNF, or both. 30-minute time points (compared to Vehicle above) are shown. Transcription factor or EP300/CTCF ChIP-peaks of either Beas-2B perturbed with Dex (B2B-DEX) or A549 cells. Numerical read distributions are not directly comparable outside of experiments due to normalization (scale on the top right of each experiment). SNPs are labeled by rsids. **C)** Ca-qtl scatterplot where each point represents a SNP that was associated with COPD or asthma in All of Us (p < 0.05), heterozygous in at least 3 ATAC-seq samples, and had minor allele frequency > 0.01 in dbGaP (Database of Genotypes and Phenotypes). Point size represents number of ATAC-seq reads containing the minor allele (normalized for total reads in library and total libraries analyzed). Red line and surrounding pink represent the genome-wide distribution and 95% confidence interval of counted allele frequency relative to population allele frequency. SNPs outside this interval = significant outliers. **D)** Top: Cartoon of the tested tRE with its two nearest genes. Bottom: Pearson correlation coefficients (PCC, x-axis) and adjusted p-values (-log10 (y-axis)) of nascent transcription between the tRE and genes within 1Mb. Horizontal line = adjusted p-value of 0.05. Colored by absolute distance of tRE to TSS of the gene. **E)** Coefficients (and standard error) of effect of tRE (vs scramble) knockout for 3 genes with adjusted p-values based on linear models.

### Focused multiomics enables the prediction of SNP function and targets in airways disease

Finally, we used multiomics data to propose epigenetic and gene regulatory functions connecting PM exposures and disease.

#### Allelic imbalance in chromatin accessibility reads predicted epigenetic mechanisms of disease

To propose SNPs and epigenetic mechanisms that link PM exposures to disease risk, we examined if chromatin accessibility read coverage showed allelic imbalance at heterozygous SNPs. We counted individual ATAC-seq reads in airway epithelial cells from heterozygous donors at each associated SNP (N=681) and computed a minor allele read count frequency, which we defined as the chromatin accessibility quantitative trait locus minor allele frequency (ca-qtl MAF). Two SNPs showed ca-qtl MAFs outside the 95% prediction interval of the genome-wide distribution and were thus defined as significant ca-qtls: rs6767543 and rs132760 (Figure 4C, Supplemental Figure CAQTLA). To determine SNPs that were functionally involved in mediating signaling responses from exposures, we also computed a Δ-ca-qtl MAF by comparing allele counts between unstimulated and exposed ATAC-seq libraries. Four of 82 tested SNPs were significant Δ-ca-qtls (Z-score > 1.96): rs11260745, rs2607940, rs6767543, and rs1864474 (Supplemental Figure CAQTLB). rs6767543 (intronic to *RARB*) met statistical criteria for both ca-qtl and Δ-ca-qtl, increased COPD risk (OR = 1.043, p-value=0.024), and was externally associated with FEV1/FVC (p-value = 2x10^-17^) and COPD (OR=1.88, p-value = 0.008) in two other studies (Huang et al., 2025). Based on ca-qtl analysis, the rs6767543 risk allele had greater chromatin accessibility than expected in unstimulated conditions but significantly decreased ca-qtl MAF and chromatin accessibility (relative corrected read count exposed/unstimulated = 0.695) following exposure. These results suggest an epigenetic mechanism where increased chromatin accessibility contributes to COPD risk at the rs6767543/*RARB* locus, which also interacts with PM exposure.

#### PRO-seq pinpoints target-genes of disease-associated SNPs

Defining genes regulated by non-coding SNPs remains a major challenge in genetic studies, so we applied and evaluated how PRO-seq data could predict the gene targets of tREs containing disease-relevant SNPs. We applied a previously established methodology that was benchmarked against CRISPR experiments, expression quantitative trait locus analyses, and three-dimensional chromatin analysis (Sigauke et al., 2025). Briefly, we computed a Pearson’s correlation coefficient (PCC) to compare nascent transcription between tREs and local genes (+/-1Mb) from both Beas-2B cells and primary smAEC samples and associated target genes with SNPs based on their resident tRE. We applied this approach to identify the plausible downstream gene mechanisms by which intergenic non-coding SNPs impact disease. We identified 152,929 significant (q < 0.001) gene-tRE pairs within 1,561,401 analyzed candidates from all Beas-2B and smAEC PRO-seq data sets analyzed in this study (n = 34 libraries). Each gene mapped to a median of 5 tREs and each tRE was connected to a median of 3 genes, consistent with estimates from lung tissue analysis in the prior publication (Sigauke et al., 2025). We further compared our results with another tool for predicting gene-tRE associations – ENCODE-rE2G (Gschwind et al., 2026). We examined only bronchial epithelial samples in ENCODE-rE2G for comparison and identified an overlap of 7,995 pairs from ENCODE-rE2G with our 152,929 significant pairs. Within these 7,995 overlaps, 93.5% were mapped to the same gene between both data sets. These experiments established to utility of applying PCC to connect genes and tREs within our nascent RNA data sets.

We then applied the PCC method to assign genes to genetic variants filtered by PM-responsive tREs and association in All of Us. We connected rs54211, which was externally associated with a reticulocyte count trait (Vuckovic et al., 2020), with *PDGFB* (PCC=0.91, q = 4.9 x 10^-11^). In contrast, the Genotype-Tissue Expression (GTEx) project had previously assigned *RPL3* (Buniello et al., 2025), which has no effect on reticulocyte biology, as the regulatory target of rs54211. Similarly, rs4989187 which had been previously associated with eosinophil counts (Verma et al., 2024), had a predicted target gene of *CYP26A1* (PCC=0.94, q = 8.4 x 10^-7^). *CYP26A1* knockout resulted in increased eosinophilic globules in a prior study (Snyder et al., 2020). Therefore, our multiomics analysis enabled the linkage of robust gene mechanisms to intergenic SNPs that were associated with disease by examining resident tREs and genes responding to PM exposures.

Finally, we experimentally verified the predicted target gene of one tRE that contained the intergenic SNP rs13243243. Based on three-dimensional chromatin conformation and expression quantitative trait locus data in lung cells, over 10 different target genes were assigned to this SNP (Supplemental Table SNPs). In Beas-2B cells, the tRE containing rs13243243 transcriptionally correlated with *TOMM7* (PCC=0.71, q = 0.004), far above all other local genes (Figure 4D). To confirm that this correlation corresponded to transcriptional regulation, we used a Cas-9 system to knock-out the tRE and measure gene expression responses to WSP exposure in Beas-2B cells (Supplemental Figure *TOMM7SNPB*). We measured gene expression of three genes – two nearest protein-coding genes (*TOMM7*, *IL6*) and one negative control gene on chromosome 15 (*CYP1A1*) – in two experiments (each N=4) of 4-hour WSP exposure using qRT-PCR (Supplemental Table qRTPCRTOMM7). A linear model of ΔΔCT based on date of experiment, genotype, and the interaction of these elements revealed that only *TOMM7* expression was significantly changed by tRE knockdown (Figure 4E, Supplemental Table lmTOMM7stat). Therefore, transcriptional correlation identified target genes for SNPs annotated to tREs and improved the definition of genes associated with both PM exposures and COPD or asthma risk.

## Discussion

Inhaled environmental exposures are a growing healthcare risk and directly contribute to respiratory disease outcomes, and we have integrated genomics methods to identify transcriptional regulatory programs that mediate this risk in airway epithelial cells. Using PRO-seq, we identified rapid-transient transcriptional responses that were shared or unique to UPM or WSP exposures in cell models. Using transcriptional networks selected for disease relevance, such as hyaluronic acid metabolism, we demonstrated that transcriptional programs identified by rapid-transient nascent RNA analysis also exhibit responses in mRNA, protein, and cellular functions. We then applied genetics and multiomics to prioritize tREs defined by nascent transcription. We showed that genetic variants found in PM-responsive tREs with nominal associations for asthma and COPD in a multi-ancestry cohort were also associated with lung function measurements, and we used summary statistics or ca-qtl analysis to further prioritize SNPs and resident tREs for functional significance. Finally, we used nascent RNA profiles to connect SNPs and tREs with target genes, and we established one association between rs13243243 and *TOMM7* using CRISPR-Cas9 genome deletion. Therefore, this study identified specific genes and mechanisms that connect PM exposures with disease risk in airway epithelial cells.

Steady-state gene expression and disease relevance supported our focus on rapid and transient transcriptional responses to particulate exposures, which are challenging to detect without nascent transcript sequencing. The strong transient response to PMs we observed might represent a non-specific initial stress response without sustained biologic impact, but rapid-transient transcription can confer changes in steady-state gene expression or prime enhancers or genes for easier accessibility when perturbed repeatedly (Vihervaara et al., 2017). We observed a clear upregulation of genes previously noted as modulating response to PM exposures, such as WNT signaling, Vitamin D metabolism, and detoxification genes (*CYP1A1* and *UGT),* and a significant proportion of early nascent transcription responses correlated with changes in mRNA at later time points (Byun et al., 2019; Guo et al., 2016; Mathyssen et al., 2019; Raslan & Yoon, 2020). Our work specifically demonstrated a rapid-transient response in hyaluronic acid metabolism that was replicated at the transcript, gene, and glycoprotein levels and reaffirmed previous associations between hyaluronic acid and COPD outcomes (Waeijen-Smit et al., 2021). Finally, the importance of rapid-transient transcription was supported by SNPs associated with COPD and asthma that were found in early response tREs and validated with disease in external multi-ancestry cohorts, such as rs12894780 and the locus on chromosome 6 containing rs28780111, rs3095340, and rs9468829. While the precise transcriptional regulatory mechanisms of these SNPs require further analysis, the association of these SNPs with disease and correlation with mRNA expression responses establish the validity of nascent transcription in defining novel and relevant rapid-transient exposure responses.

In addition to identifying a novel set of disease-relevant exposure responses, PRO-seq also connected exposures, genetics, and disease by identifying specific upstream (TF) and downstream (gene) mechanisms. For example, based on transcription factor enrichment analysis, we identified NRF factors, which are involved in antioxidant roles and mitochondrial functions, as regulators of PM exposures (Lee et al., 2021). We then demonstrated that the tRE containing the disease variant rs13243243 regulated *TOMM7*, which is also involved in mitochondrial function (Young et al., 2022). While several prior studies have indicated that mitochondrial dysfunction is a feature of COPD or asthma (Prakash et al., 2017), the precise factors that we identified improve the potential for translating this mechanism into treatment. Similarly, AHR and AHRR signaling are mediators of exposures, inflammation, and disease (Beamer & Shepherd, 2013; Coumoul et al., 2026), but variable and contradictory responses to AHR/AHRR ligands have challenged the understanding or therapeutic utilization of AHR/AHRR biology. By interrogating multiple model systems using PRO-seq, we showed that the same transcription factor – AHRR – can regulate different tREs based on cell or exposure context, and this insight may be critical to identifying a niche for AHR/AHRR-based therapy. Finally, we provided unique insight into the target genes of genetic signals, overcoming a major challenge of prior GWAS. Even where prior expression quantitative trait locus data was available, such as GTEx, we improved the assignment for genes to disease-relevant SNPs. These tools enable advanced mechanistic insight from both exposure and genetic studies, which is critical to develop novel treatments.

We applied an alternative approach to conventional GWASs by using multiomic data to power multi-ancestry analysis and focus on functional SNPs that define robust relationships between exposures and COPD or asthma. This approach leverages the observation that multi-ancestry studies are better suited to define genetic signals that are relevant to disease biology (Han et al., 2025; Shrine et al., 2023b). To overcome the statistical challenges of multi-ancestry study, we used multiomics to focus discovery and validation using genomic annotations relevant to exposures and disease biology. This approach also highlighted functional variants, which are commonly de-prioritized in favor of the statistical lead variant (Schaid et al., 2018). While the effect sizes of our high-priority SNPs were modest, we connected them with genomic and cellular functions that are biologically plausible as links between particulate exposures and COPD or asthma. For example, the genomic sites of epigenetic regulation that we proposed using ca-qtl analysis were enriched for external associations with lung function, suggesting that detailed study of these regions can define epigenetic mechanisms of disease. Using transcriptional correlation, we linked genetic signals previously unassigned to biologically relevant gene mechanisms, such as *CYP26A1* regulating eosinophilic function or *TOMM7* regulating mitochondrial function, that have direct implications in the pathogenesis and treatment of disease (Kharbili et al., 2024; Li et al., 2024; Snyder et al., 2020; Sui et al., 2024; Ying et al., 2025). Further clarification and both biological and clinical validation are still necessary to realize the objectives of this study, but candidate genes supported by exposure biology and human genetics address two main risk factors for COPD and asthma.

## Limitations of the study

Finally, our study contained notable limitations. First, nascent transcript analysis, qRT-PCR, and ELISA were performed in a limited number of samples, where donor-specific features can influence responses. To limit this bias, we excluded donors with known prior exposures, such as tobacco smoking or respiratory disease and validated genomic analysis using RNA-seq and ATAC-seq. Second, the effect sizes of our results, including SNP odds ratios and qPCR relative expression, were small. This could reflect that complex diseases are characterized by diffuse genetic signals with small effect sizes that combine with non-genetic factors to influence disease risk (Stringer et al., 2011). Our genetic analysis did not quantify interactions between each SNP and transcriptional regulation by particulate matter exposures, and including a formal interaction framework is necessary to identify true gene-by-environment effects. In addition, our model only examined a single acute exposure. Disease risk from particulate matter is likely compounded over repeated exposures and particulates are rarely inhaled in isolation. Real-world testing using exposure mixtures or advanced models are necessary to improve the relevance of our results. Therefore, even a small change in gene expression associated with a variant allele or enhancer deletion may become increasingly important over time. Despite the limitations of our study, we found transcriptional mechanisms with plausible connections to both PM exposures and asthma or COPD risk. While additional validation is necessary, specific targets have the potential to define new targeted therapies for asthma and COPD, which will be critical to mitigate the growing impact of air pollution on population health.

## Methods

### Cell Culture

Deidentified primary human small airway epithelial cells (smAECs; <2 mm bronchiole diameter) and primary human nasal airway epithelial cells (NECs) were obtained from the National Jewish Health Biobank from donors with no history of deployment or chronic lung disease (Sm36). Cells were first co-cultured with irradiated NIH/3T3 (ATCC) fibroblasts in F-medium containing 1 µM Y-27632 (APEX Bio). Cells were then passaged onto double collagen-coated transwell plates (6-well 500K cells/well; 2 plates per treatment for smAEC and 12-well, 120K/cells/well; 23 wells per treatment for NEC) and grown to confluence in fully supplemented Pneumacult medium (StemCell Technologies) containing Y-27632. Once confluent, smAEC cells were transitioned to air-liquid interface (ALI) in fully supplemented Pneumacult medium without Y-27632 and allowed to differentiate for 21 days. Once confluent, NEC cells were either treated (ALI D0) and harvested for ATAC-seq as described below or transitioned to air-liquid interface (ALI) in fully supplemented Pneumacult medium without Y-27632 and allowed to differentiate for 21 days. All cells were maintained in 5% CO2 at 37°C. For UPM treatment of smAECs, Sm36 cells differentiated for 21 days at ALI were treated with vehicle (PBS) or Urban Particulate Matter (UPM; SRM 1649b from National Institute of Standards and Technology (NIST)) at 150 ug/ml in fully supplemented Pneumacult medium applied to both apical and basal sides of membranes for 30 or 120 minutes. For treatments of NECs, cells were treated with vehicle (PBS), woodsmoke particles (WSP; 1 mg/ml) (Gupta et al., 2021) or Urban Particulate Matter (UPM; 300 ug/ml) in fully supplemented Pneumacult medium applied to both apical and basal sides of membranes for 30 or 120 minutes.

We cultured Beas-2B cells as described previously (Gupta et al., 2021, 2025). Briefly, cells were obtained from American Type Culture Collection and cultured in Dulbecco’s modified Eagle’s medium (Corning) with l-glutamine and 4.5 g/l glucose supplemented with 10% fetal bovine serum (VWR) and 1% penicillin/streptomycin.

### PRO-seq

Cells required treatment and harvest on two separate days to generate enough nuclei to perform Precision Run-on Sequencing (PRO-seq) (Gupta et al., 2021; Mahat et al., 2016; Sasse et al., 2019) on two replicates per condition [untreated control (Vehicle - VEH) vs. UPM]. For each set of replicates, Sm36 cells (small airway epithelial cells from donor 36) were seeded on 5 × 15-cm double collagen-coated plates per treatment and grown to confluence (3–4 days), then left untreated (VEH) or treated with UPM (100 µg/cm^2^) for 30 or 120 minutes. Plates were washed in 3 times in ice-cold PBS and cells were scraped in Lysis Buffer (pooling 12 wells per treatment) prior to isolation of nuclei. Nuclear run-on reactions were performed in duplicate on aliquots of 10 million nuclei in the presence of Biotin-11-CTP, then total RNA was isolated using TRIzol LS Reagent (Invitrogen). RNA was hydrolyzed with NaOH to yield fragments ∼100 bp long and nascent RNA was enriched using M-280 Streptavidin Dynabeads (Invitrogen). Sequencing adapters were ligated onto 3’ ends of nascent RNA and then a second enrichment was performed using streptavidin beads. 5’ ends were de-capped (RppH) and repaired (PNK) prior to ligation of sequencing adapters and the third and final streptavidin bead enrichment. Reverse transcription was then performed and purified cDNA libraries were amplified for sequencing in PCR reactions containing Illumina TruSeq Small RNA PCR Index Primers to differentially barcode the libraries and enable multiplexing. PCR-amplified libraries were size-selected for a range of ∼140–350 bp (average fragment length ∼250 bp) using Agencourt AMPure XP beads (Beckman Coulter) at a volume of 1X as instructed by the manufacturer. Libraries were quantified using a KAPA Library Quantification Kit for Illumina Platforms kit (KAPA Biosystems) and pooled for sequencing on an Illumina NovaSeq instrument using 150 bp reads at the University of Colorado Anschutz Medical Campus Cancer Center Genomics Shared Resource Core Facility.

### ATAC-seq

Nasal epithelial cells were washed twice with ice-cold 1X PBS and collected by scraping (pooling 3 wells/treatment) and pelleting prior to counting. Approximately 50K cells were pelleted and processed in duplicate as described in the Omni-ATAC-seq protocol published by (Corces et al., 2017)). Uniquely indexed sequencing adapters from the Nextera DNA CD Indexes Kit were added to both ends of transposed fragment libraries by PCR. Following final purification, libraries were pooled for sequencing on an Illumina NovaSeq 6000 instrument using 150 bp paired-end reads at the University of Colorado Anschutz Medical Campus Cancer Center Genomics Shared Resource Core Facility.

### qRT-PCR

smAEC cells were grown to confluence in 12-well tissue culture dishes as previously described (Gupta et al., 2022) and treated with vehicle (PBS), UPM, or WSP for 3 or 6 h. Cells were harvested in TRIzol (Life Technologies) and RNA purified by PureLink RNA Mini Kit (Life Technologies) prior to qRT–PCR, performed with normalization to *RPL19* as previously detailed(Sasse et al., 2019). Sequences of primers used for qRT–PCR analysis are provided in Supplemental Table Primers. Experiments were performed in quadruplicate with independent handling after cell seeding. That is, exposures, cell harvests, RNA purification, reverse transcription, and qPCR-based quantification were all performed independently for each of 4 wells. Experiments were repeated on separate days.

For the Cas9 knockdown, there were two experiments (N=4) on two days used, and the following statistical analysis was used. Linear models of formula ΔΔCT ∼ Genotype*Date were done for each gene using lm() in R. The Main effect was considered the Genotype coefficient (GenotypeCas9-Enh). Full results from this can be found in Supplemental Table 6. Adjusted p-values were done using the Benjamini-Hochberg approach to address for 3 genes being considered.

### ELISA

Hyaluronic Acid Enzyme-linked immunosorbent assay (ELISA). The ELISA kit was purchased from R & D Systems (DY3614). This kit detects low, medium and high molecular weight hyaluronic acid. Assay plates are coated with primary capture antibody. After sample addition, biotinylated detection antibody is added. Streptavidin-conjugated peroxidase is then added, and quantification of hyaluronic acid is performed using a colorimetric assay. Absolute quantification is calculated by comparison with a standard curve and adjusted for sample dilutions. Assays were performed in quadruplicate with independent handling after cell seeding. That is, each well was exposed independently and samples from each well were collected and analyzed separately on the same ELISA plate. Experiments were repeated twice on independent days.

### CRISPR-Cas9 Genome editing

We performed CRISPR-Cas9 genome editing to delete tREs and test tRE-gene interactions as described previously (Gupta et al., 2025). We used the Alt-R CRISPR-Cas9 system supplied and guided by IDT. Custom-designed crRNAs targeting two sites within the tRE and negative control crRNAs (IDT) were duplexed (5 min at 95°C) with tracrRNAs and then complexed with Cas9 enzyme (1:1 molar ratio of duplex to Cas9). This ribonucleic protein complex was transfected into Beas-2B cells using Lipofectamine CRISPRmax (Life Technologies) Genome editing was confirmed by PCR amplification of genomic DNA (DNeasy Blood and Tissue kit – Qiagen) before exposure to WSP and quantification of gene expression response by qRT-PCR. crRNA sequences are provided in Supplemental Table Primers. All experiments were performed in quadruplicate with independent handling after cell seeding. That is, transfection, RNA purification, reverse transcription, and qPCR-based quantification were all performed independently for each of 4 replicates during each experiment. Experiments were repeated on separate days.

### Computational Analyses

#### Genome

Unless noted otherwise, hg38 genome was used and Refseq annotations from GrCh38.p14 were used. Putative transcripts were included for enhancer annotation (whether intronic, exonic, intergenic) but not for differential expression analysis.

#### Fastq Processing, Mapping, QC (Preprocessing)

Unless otherwise noted, samtools v1.8, bedtools v2.28, python v3.6.3, bbmap v38.05, fastqc v0.11.8, hisat2 v2.1.0, preseq v2.0.3, and igvtools v2.3.75 were used. In all cases, fastq files were qc’d with FastQC, trimmed for adapters, mapped to hg38 using hisat2, and resulting SAM/BAM files qc’d before being converted to TDF format using igvtools for visualization. Scripts for these steps can be found in Preprocessing.

RNA-seq fastqs were qc’d, trimmed, and mapped to hg38 using the Nextflow pipeline https://github.com/Dowell-Lab/RNAseq-Flow (commit 6e73ba2). Fastq files from PRO-seq were qc’d, trimmed, and mapped to hg38 using the Nextflow pipeline https://github.com/Dowell-Lab/Nascent-Flow (commit 42add22). ATAC-seq fastq files were trimmed for adapters, minimum and maximum length, and minimum quality using the bbduk tool from the BBMap Suite (v38.73) with arguments “ref=adapters.fa ktrim=r qtrim=10 k=23 mink=11 hdist=1 maq=10 minlen=20 ftr=36.” Quality control was monitored both pre- and post-trim for all samples using FastQC. Trimmed reads were mapped to the human genome (hg38; downloaded from the University of California Santa Cruz genome browser on September 16, 2019, with corresponding hisat2 index files) using hisat2. Resulting SAM files were converted to sorted BAM files using samtools (v1.9). Read coverage was then normalized to reads per million mapped using a custom python script, and files were converted to TDF format using igvtools (v2.5.3) for visualization in IGV. Duplicated reads in BAMS were confined to one copy.

#### Getting enhancer coordinates from PRO-seq (Preprocessing)

Tfit v1.0 (Azofeifa & Dowell, 2017) was run for each experiment using Nextflow pipeline Bidirectional-Flow with the updates outlined in (H. A. Townsend et al., 2025) (https://github.com/Dowell-Lab/Bidirectional-Flow (branch Tfit_focus, commit cb74496)). 3’ bedgraphs were used as input into Tfit using the --tfit_3prime parameter. Consensus bidirectionals across the three perturbation experiments (WSP, UPM, ADP) were identified using muMerge v1.1.0 (https://github.com/Dowell-Lab/mumerge).

LIET-EMG (https://github.com/Dowell-Lab/LIET/tree/LIET_EMGtoo, branch LIET_EMGtoo, commit f3b1b31) was then using muMerge µs. Annotations provided to LIET included the start being the µs from muMerge, and ends being 200bp downstream. Paddings were 3kb up and downstream unless there was another enhancer mu within that region in which case the distance between enhancers was used, with a minimum of 500bp. Any cases where the 95^th^ percentile of the LIET model extended beyond the pad had paddings decreased by 200bp in the relevant direction (upstream/downstream), with a minimum pad of 300bp. As described in previous work (H. A. Townsend et al., 2025), we used a weighted average (weighted by coverage not attributed to background) of the 95^th^ percentile of the LIET-EMG model to get consensus lengths from the particulate experiments. Code for all these steps can be found in the Preprocessing section.

#### Differential expression analysis (Counting and Diff_Exp)

For RNA-seq, featurecounts (subread v1.6.2) was used with parameters -0, -s 1, -t “exon” according to the Refseq GTF. For PRO-seq, genes and bidirectionals were counted over using the Nextflow pipeline at https://github.com/Dowell-Lab/Bidir_Counting_Analysis (commit 4076721) using count type “MU_COUNTS” and the count_limit_bids of 60. This pipeline was run separately for each experiment (ADP, UPM, WSP) but using the same mumerged regions (cons_file). Briefly, this pipeline identified gene TSS bidirectionals from tREs, and addressed overlapping regions in counting (Townsend, 2025). Genes were counted over using 5’ truncated regions as detailed in (Sigauke et al., 2025; H. A. Townsend et al., 2025), with regions where bidirectionals were highly transcribed removed. For ATAC-seq, 1kb regions were used (500bp around mus). Featurecounts was again used, counting over bams with duplicated reads de-duplicated. Code and results for counting can be found in the Github under Counting.

All differential expression analysis was done using DESeq2 (v1.44.0) with PRO-seq and RNA-seq size factors calculated based on a list of housekeeping genes for the sake of consistency (size factors based on all gene counts were very comparable). For ATAC-seq analysis of enhancers, the size factors were based on DESeq2 size factors when using merged enhancer regions to ensure no double counting (non-strandedness of ATAC-seq means counts can’t be deconvoluted between overlapping regions), but the analysis was done using counts from the non-merged regions. Overenrichment analysis of GO terms was done using org.Hs.eg.db (v3.19.1) and clusterProfiler::enrichGO (v4.12.6), using all possible ontology terms, and a multiple correction adjustment using the Benjamini-Hochberg approach. Due to the large number of significantly enriched terms, we then summarized significant terms into categories. WSP and UPM PRO-seq both showed extremely robust responses, likely increasing the false positive rate. Therefore, we used more conservative adjusted p-value cutoffs (1e-10 for genes and 0.001 for enhancers) to reduce false positive calls and ensure downstream functional analyses were focused on regions with high confidence of change. General results do not change when increasing stringency of cutoffs to 1e-20. In all other cases, an adjusted p-value cutoff of 0.05 was used. Code, notebooks, and results from all differential expression analyses can be found in the Github under Diff_Exp.

#### Differential transcription factor motif enrichment (TFEA-LE)

TFEA (from Github https://github.com/Dowell-Lab/TFEA/commit 76ef85a) was used with HOCOMOCO motifs from v12. TFEA was run separately for TSS bidirectionals and tRE bidirectionals, and for each timepoint and condition (e.g. WSP 30min vs Veh). TF motif p-value cutoffs were edited as suggested by original authors to get the estimated number of TF motif instances to be between 700 and 5000 to ensure p-values were not shrunk due to simply high number of motif instances. TF Motif ZN362.H12CORE.0.P.C was not included in analysis because it had motif instance numbers above 15,000. The code for this and final p-values can be found in the Github. Final TF significant calls were done based on Townsend et al. 2025: motif instances between 600 and 10000, Fraction above background <= 0.46, GC-corrected adjusted p-value <= 0.01, and the Match background leading edge is above the plateaued enrichment leading edge but below half the number of tested enhancers.

SRF and AhR/AhRR bidirectionals were assigned by considering those enhancers within the plateaued enrichment leading-edge from TFEA-LE that also had a motif instance within 500bp of the enhancer µ. This was done for each timepoint and condition (e.g. WSP 30min vs Veh).

#### Comparing enhancer and TF calls

Enhancers and genes were assigned to different timing categories based on their response across the 30min and 120min timepoints, when available. Early rise (or fall) features were those that were significantly increasing (or decreasing) at the 30min mark compared to vehicle and decreasing (or increasing) at the 120min mark compared to 30min and were not significantly different between vehicle and 120min. Plateau rise (or fall) features were those that were significantly higher (or lower) in both 30min and 120min compared to vehicle with no significant change between 30min and 120min. Late rise (or fall) features were those that significantly increased (or decreased) expression compared to Vehicle and 30 min, but were not significantly changed at 30min compared to Vehicle. For RNA-seq, the 30min was replaced by the 2hr (earliest time point) and 120min with 4hr (latest time point).

#### caQTL Analysis

Deduplicated bam format reads from 48 unstimulated ATAC-seq (24 samples sequenced in duplicate, 15 total donors) libraries and 44 exposed ATAC-seq libraries (22 samples sequenced in duplicate, 5 total donors) were used for ca-qtl analysis. Details of each sample are provided in Supplemental Table caqtl. SNP rsid, genome coordinates, and population allele frequencies were obtained from the latest release of dbsnp156 (https://ftp.ncbi.nih.gov/snp/latest_release/VCF/GCF_000001405.40.gz) First, a consensus set of chromatin accessibility peaks was defined by applying MACS2 (Zhang et al., 2008) with a q < 1x10^-5^ threshold to each library and then using muMerge (Rubin et al., 2021) to combine all peaks. SNPs identified as “COMMON” (minor allele frequency > 0.01) were merged with this consensus peak set to identify the set of SNPs to develop the genome-wide distribution. Next, a custom python script, which is provided in github repository was used to count alleles at each SNP locus. The genomic sequence at each SNP was isolated using pysam (v0.23.3) and each allele at the SNP (A, C, G, or T) was counted. Only libraries with non-zero counts for 2 or more alleles were included (obligate heterozygotes) due to an inability to distinguish between homozygosity and inadequate read depth. The allele count was normalized to the total number of reads in the library and a final allele frequency was produced for each allele at each SNP. Next, SNPs with reads in 2 or fewer libraries were excluded. Using another custom python script, which is provided in the github repository, dbGaP population allele frequencies were extracted from the dbsnp download file for each SNP at each allele. Ultimately, allele frequencies for 69,203 were compared between expected (dbGaP) and observed (counted in ATAC-seq reads). A four-parameter logistic regression was applied to these data and a 95% prediction interval was set around the regression line to define a statistical threshold. Next, allele counts were repeated for the set of SNPs genetically associated with asthma or COPD in All of Us (n = 681) using both unstimulated (n = 48) and exposed (n = 44) libraries. Delta-ca-qtl was defined as the change in allele frequency for each allele at each SNP between unstimulated and exposed conditions. A Z-score method was applied to identify significant delta-ca-qtl SNPs based on allele frequency change values that fell outside the 95% confidence interval of the distribution of all SNPs in the testing set.

#### Genomic association analyses

We analyzed data from the All of Us Research Program, a diverse cohort including participants of European, African, Asian-American, and other ancestries. Asthma and chronic obstructive pulmonary disease (COPD) were identified from harmonized electronic health records mapped to the OMOP Common Data Model, which standardizes ICD-9/10 and SNOMED codes into unique condition concept IDs. Asthma cases were defined by ≥1 occurrence of OMOP concept IDs (317009, 40483397, 4308356, 4191479, 4125022, 46270030, 45766728, 256448, 4250128, 46270028, 4123253, 4279553, 42535716, 4309833, or 46273454). COPD cases were defined using concept IDs 25573, 4110056, and 257004. Participants with both conditions were excluded. The final analytic set included 5,843 COPD, 20,454 asthma, and 158,404 controls. SNPs with ≥90% completion and MAF ≥0.05 were retained; individuals with >10% missing genotypes were excluded. Logistic regression using SNP dosage tested COPD vs. controls and asthma vs. controls, adjusting for sex, age, pack-years, and 16 principal components. A meta-analysis combining COPD and asthma results was conducted using sample size–weighted p-values to assess shared genetic effects.

For each phenotype, linkage disequilibrium clumping was performed in PLINK 1.9 using the analyzed All of Us array-genotype sample as the LD reference. Index variants were selected at P < 0.05 and variants within 500 kb and r² ≥ 0.10 were assigned to the same clump when P < 0.05. Clumping was conducted separately for COPD and asthma. Effect-allele frequencies were calculated in the corresponding analyzed phenotype sample and aligned to the tested allele from the additive logistic-regression result.

#### Predicting Transcriptional Networks

To predict the transcription factors associated with a given tRE, we considered the presence of binding motifs, whether the TF was considered active based on global motif response (TFEA-LE), and ChIP-seq data from ENCODE. To predict the target genes of a given tRE, we considered quantitative trait loci (QTL), and nearest gene coordinates. We also recently showed that target genes show nascent transcription correlated with that of their tREs; so we also considered the genes with the highest correlation of nascent transcription across lung cell nascent sequencing samples(Gupta et al., 2021, 2022; Sasse et al., 2019; Sigauke et al., 2025). Github Repo https://github.com/Dowell-Lab/bidir_gene_pairs/tree/windowed_correlations (branch windowed_correlations, commit 32eea2ddd206c83e35c5876744ba08bc7683b0a8) was used. TPM-normalized counts were used.

Comparison with ENCODE-rE2G was performed by downloading two ENCODE accession files (ENCFF276WUK and ENCFF966SJP) and analyzing first for enhancer overlap and then for gene assignment. Positive correlations were identified as enhancers that were shared between either ENCODE data set and the candidate gene-tRE pairs and mapped to the same gene. During the preparation of this work, the authors used ChatGPT-Codex to write the custom python script used to perform this analysis. After using this tool, the authors reviewed and edited the script as needed and take full responsibility for the content of the published article. Python scripts are available in the github repository.

#### Fine mapping and candidate SNP selection

We ranked SNPs for manual annotation based on whether the meta-p-value was significant and if the odds ratio was in the same direction for both asthma and COPD. SNPs were further annotated from both external (accessed via APIs) and internal data in a code pipeline built to be reusable for later analyses. The code collecting all this data from APIs or our own datasets can be found in SNP_Analysis/. The full annotations can be found in Supplemental Table SNPs with descriptions of columns found in Supplemental Table SNP Columns. Importantly, we noticed that the web-server data was sometimes more up-to-date than that provided by the API databases. Therefore, we still encourage users to use the direct web-servers to explore a SNP of interest. For this work, APIs were accessed September 12, 2025.

##### Bidirectional Type

The bidirectional(s) in which the SNP was found for our dataset were labeled as either intergenic, exonic, intronic, Gene TSS, or LIET Gene TSS Intron/Exon/Intergenic. Gene TSS bidirectionals were first annotated according to the above Nextflow pipeline for counting. This pipeline aims to assign one active TSS bidirectional to each annotated gene isoform by choosing the bidirectional with a µ closed to the annotated TSS. We next assigned the non-TSS bidirectionals from this pipeline to intergenic, introns, exons, or gene TSS regions based on 100bp regions around the µs. Such regions were overlapped with exons, introns, and Gene TSS regions of the hg38.p14 genome using bedtools intersect with options (e.g. bedtools intersect -wo -f .51 -a ${BID} -b ${EXON}) so that the bidirectional is assigned to either introns or exons, but not both. Gene TSS regions are 1kb (500bp up and downstream of the Gene TSS). Sometimes the µ of a bidirectional is intergenic but LIET-based lengths of the bidirectional indicate that it overlaps the TSS region. We considered that these bidirectionals might be unannotated alternative start sites and therefore labeled them as LIET Gene TSS bidirectionals with extra labels of Intron/Exon/Intergenic to indicate which other region the bidirectional falls into. The code and more detailed results from this analysis can be found in Preprocessing/LIET/Get_LIET_results_bed_5.15.25.ipynb.

##### Disease Associations

SNPs associated with diseases in external analyses were noted based on GWASCatalog (Cerezo et al., 2025), ClinVar (Landrum et al., 2018), and OpenTargets (API v25.4.4) (Buniello et al., 2025). Full annotations included any variants with predicted linkage disequilibrium above 0.8 for OpenTargets and if the posterior probability was above 0.05 and absolute beta value above 0.1. The 44 SNPs considered externally associated with a phenotype were all the lead SNPs. When available, we saved the trait, beta, posterior probability (of the SNP), types of association, predicted targets of SNP, scores for the predicted target linkage, locus size (number of SNPs possibly contributing to the Beta value), and whether the lead SNP for the association is the same as the SNP of interest.

##### Molecular Effects

We first calculated the distance from the SNP to the µ of overlapping bidirectionals since the µ is predicted to estimate the transcription initiation site. SNPs overlapping TF Binding sites based on ENCODE were annotated based on the OpenTargets (API v25.4.4) (Buniello et al., 2025), with the transcription factor, and cell types and tissues in which the binding site saved. OpenTargets also overlaps variants with features annotated by ENCODE’s chromHMM (e.g. Enhancer, Genic-Enhancer, TssAFlnk, etc.)(Moore et al., 2020). We saved the regions the SNPs overlapped with, and the celltypes and tissues where the overlapping ChromHMM is found based on the OpenTargets API. Molecular effects (e.g. intron_variant, missense_variant) were saved along with the variant, predicted effect, method for prediction (e.g. Ensembl VEP), amino acid changes or distance to a footprint (e.g. TF motif instance), and predicted targets/effect scores if available again from OpenTargets API. Full annotations included any variants with predicted linkage disequilibrium above 0.8 for molecular effects and chromHMM predictions.

##### Target Genes

First, genes whose 5’ and 3’ ends were the most proximal were recorded. Second, genes with the highest correlation of transcription with the enhancer(s) were recorded based on all non-cancerous lung cells, or just Beas-2B cells or smAEC cells. The correlations of the most proximal genes were also recorded. Finally, we recorded quantitative trait loci and 3D Chromatin-based linkages (e.g. within loop, anchor-to-anchor) to genes based on the Open Targets and 3DSNPv2 APIs (Quan et al., 2022). For Open Target-based findings, full annotations included any variants with predicted linkage disequilibrium above 0.8 for these, with cases of the variant of interest being the lead SNP being noted. The tissues for the interactions, and if available, betas, posterior probabilities (of SNPs), and p-values were recorded. The transcriptional changes in particulate matter responses for all potential target genes were recorded.

#### Formal Replication using Lung function summary statistics

We downloaded summary statistics from the GWAS catalog for two accessions (GCST90244092 for FEV1 and GCST90244094 for FEV1/FVC) from the multi-ancestry cohort of a prior lung function GWAS (Shrine et al., 2023a). Next, we used the set of candidate variants with nominal p values for asthma or COPD after pruning variants by linkage disequilibrium clumps (n = 612). Among these variants, 600 were coded in the lung function summary statistics. We next removed the full 681 variants from the background data set and assembled 100,000 permutations of 600 biallelic variants matched on minor allele frequency and calculated the average -log_10_ P value for each permutation as well as our candidate set. We calculated an empirical p value based on comparison with the null/background distribution generated from the 100,000 permutations and applied the Benjamini-Hochberg method for adjusting for multiple testing for multiple phenotypes. During the preparation of this work, the authors used ChatGPT-Codex to write the custom python script used to perform this analysis. After using this tool, the authors reviewed and edited the script as needed and take full responsibility for the content of the published article. Python scripts for this analysis are included in the github repository.

## Resource Availability

### Lead Contact

Arnav Gupta –

### Materials Availability

All resources presented in this manuscript with exceptions for commercially purchased or proprietary materials can be made available by request to the lead contact. Primary cells from multiple donors require permission from the National Jewish Health Cell Core due to Institutional Review Board policy.

### Data and Code Availability

DEX-TNF Beas2B GRO-seq (GSE124916), WSP Beas2B RNA-seq (GSE283113), WSP Beas2B PRO-seq (GSE167372), WSP/UPM NEC ATAC-seq (GSE327419), UPM smAEC PRO-seq (GSE326905), ADP smAEC PRO-seq (GSE201150). ENCODE experiments and files, all in A549, include ENCSR977FEF (PRDM1 Experiment), ENCFF096PTH (EP300), ENCFF229ULU (CTCF), ENCFF835PPK (YY1), ENCFF618CNL (JUN), ENCFF229ULU (TP53), ENCFF271WHK (ETS1), ENCFF343BIK (GABPA), ENCFF232TVP (FOSL2), ENCFF169QCM (ELF1). For ca-QTL analysis, the following GEO datasets were used: GSE201149, GSE167370, three unstimulated, healthy donors in GSE283113, and GSE327935.

All results/data used for analysis not directly found in this work can be found at Zenodo: (H. Townsend et al., 2026). All code can be found at the Github repository https://github.com/Hope2925/UPM-Particulate-SNP.

## Contributions (based on CRedIT)

HAT: Data curation, Formal analysis, Methodology, Software, Validation, Visualization, Writing – original draft, Writing – review & editing

SKS: Conceptualization, Data curation, Investigation (PRO-seq & ATAC-seq), Methodology, Writing – review & editing

SL: Formal analysis (Genomics association), Methodology (Genomics association), Writing – review & editing

A Gerber: Conceptualization, Supervision, Writing – review & editing

RDD: Conceptualization, Methodology, Supervision, Writing – review & editing

A Gupta: Conceptualization, Data curation, Formal analysis (caQTL), Investigation, Methodology (caQTL), Validation (CRISPR-Cas9), Writing – original draft, Writing – review & editing

## Acknowledgements/Funding

Thank you to Dr. Fabienne Gally for her advice on this project. Thank you to the Biofrontiers Institute of Technology team for ensuring the computational work ran smoothly.

National Institutes of Environmental Health Sciences 1K08ES034820 (PI: Gupta)

Support for this study was also provided by the National Emphysema Foundation to Arnav Gupta.

This work was supported by NHLBI grants U01 HL089897 and U01 HL089856 and by NIH contract 75N92023D00011. The COPDGene study (NCT00608764) has also been supported by the COPD Foundation through contributions made to an Industry Advisory Committee that has included AstraZeneca, Bayer Pharmaceuticals, Boehringer-Ingelheim, Bristol Myers Squibb, Genentech, GlaxoSmithKline, Johnson & Johnson, Novartis, Pfizer, Regeneron, and Sunovion.

## Declaration of Interests

Dr. Dowell is a founder of Arpeggio Biosciences. Dr. Dowell is on a patent for inferring TF activity from eRNA activity (PCT/US2018/0182330). The remaining authors declare no competing interests.

## Supplemental Information

Document Supplemental_figures.pdf – Contains Supplemental figures reference by individual names

Data Supplemental_tables.xlsx – Contains Supplemental Tables 1 – 15.

## Notes

### Author Declarations

This study ysed only openly available human data contained in the database of genotypes and phenotypes (dbGaP), the All of Us research Program, or the Genetic Epidemiology of COPD study (data available in dbGaP).

### Summary of Updates

Given the central importance of the PCC method in our study, we have improved the statistical rigor of this analysis, including comparison with prior publication, systematic analysis of gene-enhancer pairs in our study, benchmarking against ENCODE-rE2G, and estimation of false discovery rate using the Benjamini-Hochberg method. We have performed a new statistical replication analysis to support the use of the genetic variants as a filter for transcribed regulatory elements. We used summary statistics from a lung function GWAS to analyze our prioritized variants in bulk and filter for specific variants.

## References

Azofeifa, J. G., & Dowell, R. D. (2017). A generative model for the behavior of RNA polymerase. Bioinformatics, 33(2), 227–234. 10.1093/bioinformatics/btw599

Beamer, C. A., & Shepherd, D. M. (2013). ROLE OF THE ARYL HYDROCARBON RECEPTOR (AHR) IN LUNG INFLAMMATION. Seminars in Immunopathology, 35(6), 10.1007/s00281-013-0391-0397. https://doi.org/10.1007/s00281-013-0391-7

Buniello, A., Suveges, D., Cruz-Castillo, C., Llinares, M. B., Cornu, H., Lopez, I., Tsukanov, K., Roldán-Romero, J. M., Mehta, C., Fumis, L., McNeill, G., Hayhurst, J. D., Martinez Osorio, R. E., Barkhordari, E., Ferrer, J., Carmona, M., Uniyal, P., Falaguera, M. J., Rusina, P., … Ochoa, D. (2025). Open Targets Platform: Facilitating therapeutic hypotheses building in drug discovery. Nucleic Acids Research, 53(D1), D1467–D1475. 10.1093/nar/gkae1128

Byun, J., Song, B., Lee, K., Kim, B., Hwang, H. W., Ok, M.-R., Jeon, H., Lee, K., Baek, S.-K., Kim, S.-H., Oh, S. J., & Kim, T. H. (2019). Identification of urban particulate matter-induced disruption of human respiratory mucosa integrity using whole transcriptome analysis and organ-on-a chip. Journal of Biological Engineering, 13(1), 88. 10.1186/s13036-019-0219-7

Campo, G. M., Avenoso, A., D’Ascola, A., Prestipino, V., Scuruchi, M., Nastasi, G., Calatroni, A., & Campo, S. (2012). Hyaluronan differently modulates TLR-4 and the inflammatory response in mouse chondrocytes. BioFactors, 38(1), 69–76. 10.1002/biof.202

Centers for Disease Control and Prevention. National Center for Health Statistics. Behavioral Risk Factor Surveillance System, 2023. Calculations by the American Lung Association Research and Program Services Division using SPSS software. (n.d.). [Dataset].

Cerezo, M., Sollis, E., Ji, Y., Lewis, E., Abid, A., Bircan, K. O., Hall, P., Hayhurst, J., John, S., Mosaku, A., Ramachandran, S., Foreman, A., Ibrahim, A., McLaughlin, J., Pendlington, Z., Stefancsik, R., Lambert, S. A., McMahon, A., Morales, J., … Harris, L. W. (2025). The NHGRI-EBI GWAS Catalog: Standards for reusability, sustainability and diversity. Nucleic Acids Research, 53(D1), D998–D1005. 10.1093/nar/gkae1070

Chen, X.-F., Guo, M.-R., Duan, Y.-Y., Jiang, F., Wu, H., Dong, S.-S., Zhou, X.-R., Thynn, H. N., Liu, C.-C., Zhang, L., Guo, Y., & Yang, T.-L. (2020). Multiomics dissection of molecular regulatory mechanisms underlying autoimmune-associated noncoding SNPs. JCI Insight, 5(17), e136477. 10.1172/jci.insight.136477

Cho, M. H., Hobbs, B. D., & Silverman, E. K. (2022a). Genetics of chronic obstructive pulmonary disease: Understanding the pathobiology and heterogeneity of a complex disorder. The Lancet Respiratory Medicine, 10(5), 485–496. 10.1016/S2213-2600(21)00510-5

Cho, M. H., Hobbs, B. D., & Silverman, E. K. (2022b). The Genetics of COPD. The Lancet. Respiratory Medicine, 10(5), 485–496. 10.1016/S2213-2600(21)00510-5

Corces, M. R., Trevino, A. E., Hamilton, E. G., Greenside, P. G., Sinnott-Armstrong, N. A., Vesuna, S., Satpathy, A. T., Rubin, A. J., Montine, K. S., Wu, B., Kathiria, A., Cho, S. W., Mumbach, M. R., Carter, A. C., Kasowski, M., Orloff, L. A., Risca, V. I., Kundaje, A., Khavari, P. A., … Chang, H. Y. (2017). An improved ATAC-seq protocol reduces background and enables interrogation of frozen tissues. Nature Methods, 14(10), 959– 962. 10.1038/nmeth.4396

Coumoul, X., Barouki, R., Esser, C., Haarmann-Stemmann, T., Lawrence, B. P., Lehmann, J., Moura-Alves, P., Murray, I. A., Opitz, C. A., Perdew, G. H., & Bourguet, W. (2026). The aryl hydrocarbon receptor: Structure, signaling, physiology and pathology. Signal Transduction and Targeted Therapy, 11, 20. 10.1038/s41392-025-02500-8

Cui, C., Yang, R., Chen, H., Li, D., Sun, X., Wang, Y., & Pan, Q. (2025). Air toxins disorder the NF-kB Pathway leads to immune disorders and immune diseases in the human health. Ecotoxicology and Environmental Safety, 302, 118474. 10.1016/j.ecoenv.2025.118474

Foundation, F. S. (2022). First Street Foundation Aggregated Wildfire Risk Summary Statistics Version 1.0 [Dataset]. 10.5281/zenodo.6564731

Galdi, F., Pedone, C., McGee, C. A., George, M., Rice, A. B., Hussain, S. S., Vijaykumar, K., Boitet, E. R., Tearney, G. J., McGrath, J. A., Brown, A. R., Rowe, S. M., Incalzi, R. A., & Garantziotis, S. (2021). Inhaled high molecular weight hyaluronan ameliorates respiratory failure in acute COPD exacerbation: A pilot study. Respiratory Research, 22, 30. 10.1186/s12931-020-01610-x

Giammona, A., Gervasoni, C., Di Iulio, G., Sirignano, C., Listrani, S., Rinaldi, M., Canepari, S., Lo Dico, A., Costabile, F., & Bertoli, G. (2025). Cellular responses of human bronchial epithelial cells following short-term exposure to oxidative particulate matter. Environmental Toxicology and Pharmacology, 120, 104853. 10.1016/j.etap.2025.104853

Gschwind, A. R., Mualim, K. S., Karbalayghareh, A., Sheth, M. U., Dey, K. K., Jagoda, E., Nurtdinov, R. N., Xi, W., Tan, A. S., Galante, J., Jones, H., Ma, X. R., Yao, D., Amgalan, D., Ray, J., Munger, C. J., Nasser, J., Avsec, Ž., James, B. T., … Engreitz, J. M. (2026). An encyclopedia of human enhancer–gene regulatory interactions. Nature, 657(8130), 179–190. 10.1038/s41586-026-10781-4

Gu, X.-J., Su, W.-M., Dou, M., Jiang, Z., Duan, Q.-Q., Yin, K.-F., Cao, B., Wang, Y., Li, G.-B., & Chen, Y.-P. (2023). Expanding causal genes for Parkinson’s disease via multi-omics analysis. Npj Parkinson’s Disease, 9(1), Article 1. 10.1038/s41531-023-00591-0

Guo, L., Wang, T., Wu, Y., Yuan, Z., Dong, J., Li, X., An, J., Liao, Z., Zhang, X., Xu, D., & Wen, F.-Q. (2016). WNT/β-catenin signaling regulates cigarette smoke-induced airway inflammation via the PPARδ/p38 pathway. Laboratory Investigation, 96(2), 218–229. 10.1038/labinvest.2015.101

Gupta, A., Dahlin, A., Macario, A., Gally, F., Weaver, M. R., Guarino, S., Kahn, L., Sanford, L., Gruca, M. A., Cho, M. H., Dowell, R. D., Weiss, S. T., Sasse, S. K., & Gerber, A. N. (2025). Functional Variant Discovery Identifies a Novel Genetic Link Between SPRY2, Wood Smoke, and Asthma. American Journal of Respiratory Cell and Molecular Biology, rcmb.2024-0587OC. 10.1165/rcmb.2024-0587OC

Gupta, A., Sasse, S. K., Berman, R., Gruca, M. A., Dowell, R. D., Chu, H. W., Downey, G. P., & Gerber, A. N. (2022a). Integrated genomics approaches identify transcriptional mediators and epigenetic responses to Afghan desert particulate matter in small airway epithelial cells. Physiological Genomics, 54(10), 389–401. 10.1152/physiolgenomics.00063.2022

Gupta, A., Sasse, S. K., Berman, R., Gruca, M. A., Dowell, R. D., Chu, H. W., Downey, G. P., & Gerber, A. N. (2022b). Integrated genomics approaches identify transcriptional mediators and epigenetic responses to Afghan desert particulate matter in small airway epithelial cells. Physiological Genomics, 54(10), 389–401. 10.1152/physiolgenomics.00063.2022

Gupta, A., Sasse, S. K., Gruca, M. A., Sanford, L., Dowell, R. D., & Gerber, A. N. (2021a). Deconvolution of multiplexed transcriptional responses to wood smoke particles defines rapid aryl hydrocarbon receptor signaling dynamics. Journal of Biological Chemistry, 297(4). 10.1016/j.jbc.2021.101147

Gupta, A., Sasse, S. K., Gruca, M. A., Sanford, L., Dowell, R. D., & Gerber, A. N. (2021b). Deconvolution of multiplexed transcriptional responses to wood smoke particles defines rapid aryl hydrocarbon receptor signaling dynamics. The Journal of Biological Chemistry, 297(4), 101147. 10.1016/j.jbc.2021.101147

Han, A. L., Sands, C. F., Matelska, D., Butts, J. C., Ravanmehr, V., Hu, F., Villavicencio Gonzalez, E., Katsanis, N., Bustamante, C. D., Wang, Q., Petrovski, S., Vitsios, D., & Dhindsa, R. S. (2025). Diverse ancestral representation improves genetic intolerance metrics. Nature Communications, 16(1), 2648. 10.1038/s41467-025-57885-5

Huang, L., Xu, W., Fu, Y., Yang, Z., Mo, R., Ding, Y., & Xie, T. (2025). RARB genetic variants might contribute to the risk of chronic obstructive pulmonary disease based on a case-control study. Annals of Medicine, 57(1), 2445195. 10.1080/07853890.2024.2445195

Kharbili, M. E., Sasse, S. K., Sanford, L., Jacobson, S., Aviszus, K., Gupta, A., Guo, C., Majka, S. M., Dowell, R. D., Gerber, A. N., Bowler, R. P., & Gally, F. (2024). Noncoding SNPs decrease expression of FABP5 during COPD exacerbations. The Journal of Clinical Investigation, 134(3). 10.1172/JCI175626

Kim, G.-D., Shin, D.-U., Song, H.-J., Lim, K. M., Eom, J.-E., Lim, E. Y., Kim, Y. I., Song, J. H., Kim, H.-J., Lee, S.-Y., & Shin, H. S. (2024). Analysis of particulate matter-induced alteration of genes and related signaling pathways in the respiratory system. Ecotoxicology and Environmental Safety, 281, 116637. 10.1016/j.ecoenv.2024.116637

Kim, N., Han, D. H., Suh, M.-W., Lee, J. H., Oh, S.-H., & Park, M. K. (2019). Effect of lipopolysaccharide on diesel exhaust particle-induced junctional dysfunction in primary human nasal epithelial cells. Environmental Pollution, 248, 736–742. 10.1016/j.envpol.2019.02.082

Landrum, M. J., Lee, J. M., Benson, M., Brown, G. R., Chao, C., Chitipiralla, S., Gu, B., Hart, J., Hoffman, D., Jang, W., Karapetyan, K., Katz, K., Liu, C., Maddipatla, Z., Malheiro, A., McDaniel, K., Ovetsky, M., Riley, G., Zhou, G., … Maglott, D. R. (2018). ClinVar: Improving access to variant interpretations and supporting evidence. Nucleic Acids Research, 46(D1), D1062–D1067. 10.1093/nar/gkx1153

Lee, J., Jang, J., Park, S.-M., & Yang, S.-R. (2021). An Update on the Role of Nrf2 in Respiratory Disease: Molecular Mechanisms and Therapeutic Approaches. International Journal of Molecular Sciences, 22(16), 8406. 10.3390/ijms22168406

Li, C.-L., Liu, J.-F., & Liu, S.-F. (2024). Mitochondrial Dysfunction in Chronic Obstructive Pulmonary Disease: Unraveling the Molecular Nexus. Biomedicines, 12(4), 814. 10.3390/biomedicines12040814

Li, Y.-J., Kawada, T., & Azuma, A. (2013). Nrf2 Is a Protective Factor against Oxidative Stresses Induced by Diesel Exhaust Particle in Allergic Asthma. Oxidative Medicine and Cellular Longevity, 2013, 323607. 10.1155/2013/323607

Mahat, D. B., Kwak, H., Booth, G. T., Jonkers, I. H., Danko, C. G., Patel, R. K., Waters, C. T., Munson, K., Core, L. J., & Lis, J. T. (2016). Base-Pair Resolution Genome-Wide Mapping Of Active RNA polymerases using Precision Nuclear Run-On (PRO-seq). Nature Protocols, 11(8), 1455. 10.1038/nprot.2016.086

Mathyssen, C., Serré, J., Sacreas, A., Everaerts, S., Maes, K., Verleden, S., Verlinden, L., Verstuyf, A., Pilette, C., Gayan-Ramirez, G., Vanaudenaerde, B., & Janssens, W. (2019). Vitamin D Modulates the Response of Bronchial Epithelial Cells Exposed to Cigarette Smoke Extract. Nutrients, 11(9), 2138. 10.3390/nu11092138

Mizuno, S., Ishizaki, T., Kadowaki, M., Akai, M., Shiozaki, K., Iguchi, M., Oikawa, T., Nakagawa, K., Osanai, K., Toga, H., Gomez-Arroyo, J., Kraskauskas, D., Cool, C. D., Bogaard, H. J., & Voelkel, N. F. (2017). *P53* Signaling Pathway Polymorphisms Associated With Emphysematous Changes in Patients With COPD. Chest, 152(1), 58–69. 10.1016/j.chest.2017.03.012

Moore, J. E., Purcaro, M. J., Pratt, H. E., Epstein, C. B., Shoresh, N., Adrian, J., Kawli, T., Davis, C. A., Dobin, A., Kaul, R., Halow, J., Van Nostrand, E. L., Freese, P., Gorkin, D. U., Shen, Y., He, Y., Mackiewicz, M., Pauli-Behn, F., Williams, B. A., … Weng, Z. (2020). Expanded encyclopaedias of DNA elements in the human and mouse genomes. Nature, 583(7818), Article 7818. 10.1038/s41586-020-2493-4

Papakonstantinou, E., Klagas, I., Miglino, N., Karakiulakis, G., Tamm, M., & Roth, M. (2014). Cigarette smoke alters hyaluronic acid homeostasis in primary human lung fibroblasts. European Respiratory Journal, 38(Suppl 55). 10.1183/13993003/erj.38.Suppl_55.p758

Petrigni, G., & Allegra, L. (2006). Aerosolised hyaluronic acid prevents exercise-induced bronchoconstriction, suggesting novel hypotheses on the correction of matrix defects in asthma. Pulmonary Pharmacology & Therapeutics, 19(3), 166–171. 10.1016/j.pupt.2005.03.002

Prakash, Y. S., Pabelick, C. M., & Sieck, G. C. (2017). Mitochondrial Dysfunction in Airway Disease. Chest, 152(3), 618–626. 10.1016/j.chest.2017.03.020

Proestakis, E., Papachristopoulou, K., Georgiou, T., Chatoutsidou, S. E., Lazaridis, M., Gkikas, A., Fountoulakis, I., Tsikoudi, I., Petrakis, M. P., & Amiridis, V. (2025). Atmospheric dust and air quality over large-cities and megacities of the world. Atmospheric Chemistry and Physics, 25(21), 14777–14823. 10.5194/acp-25-14777-2025

Quan, C., Ping, J., Lu, H., Zhou, G., & Lu, Y. (2022). 3DSNP 2.0: Update and expansion of the noncoding genomic variant annotation database. Nucleic Acids Research, 50(D1), D950– D955. 10.1093/nar/gkab1008

Raslan, A. A., & Yoon, J. K. (2020). WNT Signaling in Lung Repair and Regeneration. Molecules and Cells, 43(9), 774–783. 10.14348/molcells.2020.0059

Rubin, J. D., Stanley, J. T., Sigauke, R. F., Levandowski, C. B., Maas, Z. L., Westfall, J., Taatjes, D. J., & Dowell, R. D. (2021). Transcription factor enrichment analysis (TFEA) quantifies the activity of multiple transcription factors from a single experiment. Communications Biology, 4(1), Article 1. 10.1038/s42003-021-02153-7

Sagiv, A., Bar-Shai, A., Levi, N., Hatzav, M., Zada, L., Ovadya, Y., Roitman, L., Manella, G., Regev, O., Majewska, J., Vadai, E., Eilam, R., Feigelson, S. W., Tsoory, M., Tauc, M., Alon, R., & Krizhanovsky, V. (2018). P53 in Bronchial Club Cells Facilitates Chronic Lung Inflammation by Promoting Senescence. Cell Reports, 22(13), 3468–3479. 10.1016/j.celrep.2018.03.009

Sasse, S. K., Dahlin, A., Sanford, L., Gruca, M. A., Gupta, A., Gally, F., Wu, A. C., Iribarren, C., Dowell, R. D., Weiss, S. T., & Gerber, A. N. (2025). Enhancer RNA transcription pinpoints functional genetic variants linked to asthma. Nature Communications, 16(1), 2750. 10.1038/s41467-025-57693-x

Sasse, S. K., Gruca, M., Allen, M. A., Kadiyala, V., Song, T., Gally, F., Gupta, A., Pufall, M. A., Dowell, R. D., & Gerber, A. N. (2019). Nascent transcript analysis of glucocorticoid crosstalk with TNF defines primary and cooperative inflammatory repression. Genome Research, 29(11), 1753–1765. 10.1101/gr.248187.119

Sato, S., Miyazaki, M., Fukuda, S., Mizutani, Y., Mizukami, Y., Higashiyama, S., & Inoue, S. (2023). Human TMEM2 is not a catalytic hyaluronidase, but a regulator of hyaluronan metabolism via HYBID (KIAA1199/CEMIP) and HAS2 expression. Journal of Biological Chemistry, 299(6). 10.1016/j.jbc.2023.104826

Sato, S., Mizutani, Y., Yoshino, Y., Masuda, M., Miyazaki, M., Hara, H., & Inoue, S. (2021). Pro-inflammatory cytokines suppress HYBID (hyaluronan (HA)-binding protein involved in HA depolymerization/KIAA1199/CEMIP)-mediated HA metabolism in human skin fibroblasts. Biochemical and Biophysical Research Communications, 539, 77–82. 10.1016/j.bbrc.2020.12.082

Schaid, D. J., Chen, W., & Larson, N. B. (2018). From genome-wide associations to candidate causal variants by statistical fine-mapping. Nature Reviews Genetics, 19(8), Article 8. 10.1038/s41576-018-0016-z

Shrine, N., Izquierdo, A. G., Chen, J., Packer, R., Hall, R. J., Guyatt, A. L., Batini, C., Thompson, R. J., Pavuluri, C., Malik, V., Hobbs, B. D., Moll, M., Kim, W., Tal-Singer, R., Bakke, P., Fawcett, K. A., John, C., Coley, K., Piga, N. N., … Tobin, M. D. (2023a). Multi-ancestry genome-wide association analyses improve resolution of genes and pathways influencing lung function and chronic obstructive pulmonary disease risk. Nature Genetics, 55(3), 410–422. 10.1038/s41588-023-01314-0

Shrine, N., Izquierdo, A. G., Chen, J., Packer, R., Hall, R. J., Guyatt, A. L., Batini, C., Thompson, R. J., Pavuluri, C., Malik, V., Hobbs, B. D., Moll, M., Kim, W., Tal-Singer, R., Bakke, P., Fawcett, K. A., John, C., Coley, K., Piga, N. N., … Tobin, M. D. (2023b). Multi-ancestry genome-wide association analyses improve resolution of genes and pathways influencing lung function and chronic obstructive pulmonary disease risk. Nature Genetics, 55(3), 410–422. 10.1038/s41588-023-01314-0

Sigauke, R. F., Sanford, L., Maas, Z. L., Jones, T., Stanley, J. T., Townsend, H. A., Allen, M. A., & Dowell, R. D. (2025). Atlas of nascent RNA transcripts reveals tissue-specific enhancer to gene linkages. BMC Genomics, 26(1), 406. 10.1186/s12864-025-11568-z

Sin, D. D., Doiron, D., Agusti, A., Anzueto, A., Barnes, P. J., Celli, B. R., Criner, G. J., Halpin, D., Han, M. K., Martinez, F. J., Oca, M. M. de, Papi, A., Pavord, I., Roche, N., Singh, D., Stockley, R., Varlera, M. V. L., Wedzicha, J., Vogelmeier, C., & Bourbeau, J. (2023). Air pollution and COPD: GOLD 2023 committee report. European Respiratory Journal, 61(5). 10.1183/13993003.02469-2022

Snyder, J. M., Zhong, G., Hogarth, C., Huang, W., Topping, T., LaFrance, J., Palau, L., Czuba, L. C., Griswold, M., Ghiaur, G., & Isoherranen, N. (2020). Knockout of Cyp26a1 and Cyp26b1 during postnatal life causes reduced lifespan, dermatitis, splenomegaly, and systemic inflammation in mice. The FASEB Journal, 34(12), 15788–15804. 10.1096/fj.202001734R

Soberanes, S., Panduri, V., Mutlu, G. M., Ghio, A., Bundinger, G. R. S., & Kamp, D. W. (2006). P53 Mediates Particulate Matter–induced Alveolar Epithelial Cell Mitochondria-regulated Apoptosis. American Journal of Respiratory and Critical Care Medicine, 174(11), 1229–1238. 10.1164/rccm.200602-203OC

Stringer, S., Wray, N. R., Kahn, R. S., & Derks, E. M. (2011). Underestimated Effect Sizes in GWAS: Fundamental Limitations of Single SNP Analysis for Dichotomous Phenotypes. PLOS ONE, 6(11), e27964. 10.1371/journal.pone.0027964

Sui, J., Xiao, H., Mbaekwe, U., Ting, N.-C., Murday, K., Hu, Q., Gregory, A. D., Kapellos, T. S., Yildirim, A. Ö., Königshoff, M., Zhang, Y., Sciurba, F., Das, J., & Kliment, C. R. (2024). Interpretable machine learning uncovers epithelial transcriptional rewiring and a role for Gelsolin in COPD. JCI Insight, 9(21). 10.1172/jci.insight.180239

The All of Us Research Program Investigators. (2019). The “All of Us” Research Program. New England Journal of Medicine, 381(7), 668–676. 10.1056/NEJMsr1809937

Townsend, H. A., Stanley, J. T., Allen, M. A., & Dowell, R. D. (2025). Improving confidence of differential transcription calls in enhancers (p. 2025.09.12.675852). bioRxiv. 10.1101/2025.09.12.675852

Townsend, H. A., Stanley, J. T., Allen, M. A., & Dowell, R. D. (2026). Improving calls of differentially transcribed enhancers and their upstream regulators. *Bioinformatics Advances*, vbag162. 10.1093/bioadv/vbag162

Townsend, H., Dowell, R., Gupta, A., Sasse, S., Gerber, A., & Liao, S.-Y. (2026). Air pollutant multiomics improves functional annotation of SNPs associated with lung disease. 10.5281/zenodo.20695133

Verma, A., Huffman, J. E., Rodriguez, A., Conery, M., Liu, M., Ho, Y.-L., Kim, Y., Heise, D. A., Guare, L., Panickan, V. A., Garcon, H., Linares, F., Costa, L., Goethert, I., Tipton, R., Honerlaw, J., Davies, L., Whitbourne, S., Cohen, J., … Liao, K. P. (2024). Diversity and scale: Genetic architecture of 2068 traits in the VA Million Veteran Program. Science, 385(6706), eadj1182. 10.1126/science.adj1182

Vihervaara, A., Mahat, D. B., Guertin, M. J., Chu, T., Danko, C. G., Lis, J. T., & Sistonen, L. (2017). Transcriptional response to stress is pre-wired by promoter and enhancer architecture. Nature Communications, 8(1), 255. 10.1038/s41467-017-00151-0

Vuckovic, D., Bao, E. L., Akbari, P., Lareau, C. A., Mousas, A., Jiang, T., Chen, M.-H., Raffield, L. M., Tardaguila, M., Huffman, J. E., Ritchie, S. C., Megy, K., Ponstingl, H., Penkett, C. J., Albers, P. K., Wigdor, E. M., Sakaue, S., Moscati, A., Manansala, R., … Soranzo, N. (2020). The Polygenic and Monogenic Basis of Blood Traits and Diseases. Cell, 182(5), 1214–1231.e11. 10.1016/j.cell.2020.08.008

Waeijen-Smit, K., Reynaert, N. L., Beijers, R. J. H. C. G., Houben-Wilke, S., Simons, S. O., Spruit, M. A., & Franssen, F. M. E. (2021). Alterations in plasma hyaluronic acid in patients with clinically stable COPD versus (non)smoking controls. Scientific Reports, 11(1), 15883. 10.1038/s41598-021-95030-6

Wan, M., Wang, C., Cui, J., Xia, Q., & Zhang, L. (2024). Silencing KLF6 Alleviates Cigarette Smoke Extract-Induced Mitochondrial Dysfunction in Bronchial Epithelial Cells by SIRT4 Upregulation. International Journal of Chronic Obstructive Pulmonary Disease, 19, 815–828. 10.2147/COPD.S451264

Watanabe, K., Taskesen, E., van Bochoven, A., & Posthuma, D. (2017). Functional mapping and annotation of genetic associations with FUMA. Nature Communications, 8(1), Article 1. 10.1038/s41467-017-01261-5

Ying, H., Wu, X., Jia, X., Yang, Q., Liu, H., Zhao, H., Chen, Z., Xu, M., Wang, T., Li, M., Zhao, Z., Zheng, R., Wang, S., Lin, H., Xu, Y., Lu, J., Wang, W., Ning, G., Zheng, J., & Bi, Y. (2025). Single-cell transcriptome-wide Mendelian randomization and colocalization reveals immune-mediated regulatory mechanisms and drug targets for COVID-19. eBioMedicine, 113. 10.1016/j.ebiom.2025.105596

Young, C., Batkovskyte, D., Kitamura, M., Shvedova, M., Mihara, Y., Akiba, J., Zhou, W., Hammarsjö, A., Nishimura, G., Yatsuga, S., Grigelioniene, G., & Kobayashi, T. (2022). A hypomorphic variant in the translocase of the outer mitochondrial membrane complex subunit TOMM7 causes short stature and developmental delay. Human Genetics and Genomics Advances, 4(1), 100148. 10.1016/j.xhgg.2022.100148

Zhang, Y., Liu, T., Meyer, C. A., Eeckhoute, J., Johnson, D. S., Bernstein, B. E., Nusbaum, C., Myers, R. M., Brown, M., Li, W., & Liu, X. S. (2008). Model-based Analysis of ChIP-Seq (MACS). Genome Biology, 9(9), R137. 10.1186/gb-2008-9-9-r137

Zhou, J. J., Cho, M. H., Castaldi, P. J., Hersh, C. P., Silverman, E. K., & Laird, N. M. (2013). Heritability of Chronic Obstructive Pulmonary Disease and Related Phenotypes in Smokers. American Journal of Respiratory and Critical Care Medicine, 188(8), 941–947. 10.1164/rccm.201302-0263OC

Zhou, W., Tian, D., He, J., Wang, Y., Zhang, L., Cui, L., jia, L., Zhang, L., Li, L., Shu, Y., Yu, S., Zhao, J., Yuan, X., & Peng, S. (2016). Repeated PM2.5 exposure inhibits BEAS-2B cell P53 expression through ROS-Akt-DNMT3B pathway-mediated promoter hypermethylation. Oncotarget, 7(15), 20691–20703. 10.18632/oncotarget.7842

Zou, K., & Zeng, Z. (2023). Role of early growth response 1 in inflammation-associated lung diseases. American Journal of Physiology - Lung Cellular and Molecular Physiology, 325(2), L143–L154. 10.1152/ajplung.00413.2022

