## Supplemental figures for "Air pollutant multiomics improves functional annotation of SNPs associated with lung disease"

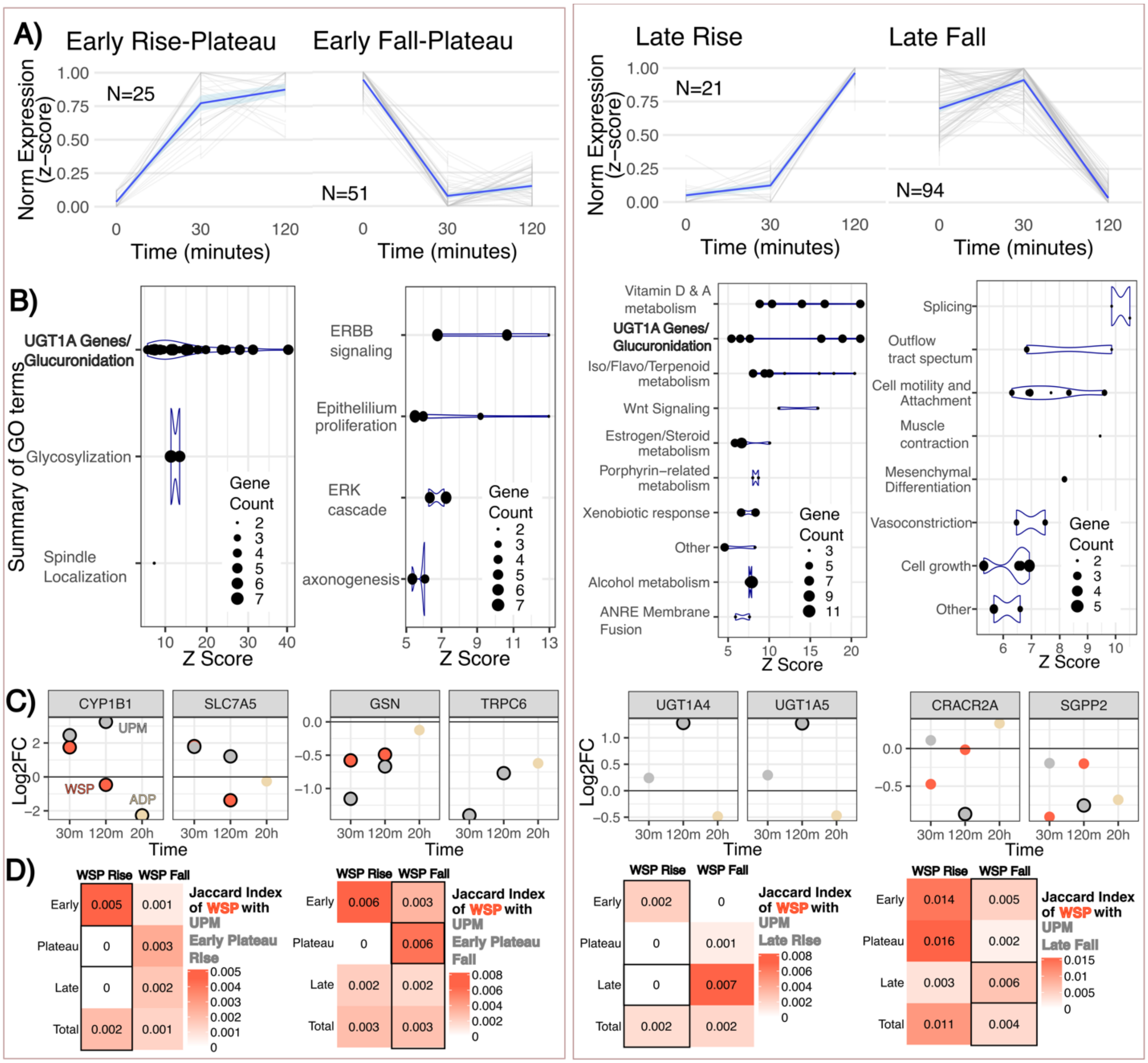


Supplemental Figure 1**. Plateau and Late Response Genes.** ***A)*** *Line plots of the mean (blue, with light blue 95% confidence interval) and individual (grey) normalized transcriptional changes of genes statistically significant (adjusted-p-value < 1x10*$^{-10}$*) matching different timeline categories (details in Methods). Norm Expression (z-score) refers to normalized counts min-max scaled. 0 minutes refers to Vehicle response.* ***B)*** *Summary of Gene Ontology (GO) terms of the matching genes (exact terms and grouping found in Zenodo) where each term is a GO term called significant with the size of the dot corresponding to the number of significant genes matching the GO term.* ***C)*** *Log2 fold change of normalized counts of the genes with the greatest statistical significance in change for UPM among three different perturbations (UPM, wood smoke particles (WSP), or Afghan dust particles (ADP)) compared to vehicle at their available timepoints. Cases where the change had an adjusted p-value less than 1x10*$^{-6}$ *were outlined in*

*black. UGT1A4 and UGT1A5 were not significantly transcribed in WSP conditions.* ***D).*** *Jaccard index of significant UPM genes for the timing category across all timeline categories of WSP responsive genes (adjusted-p-value < $1x10^{-10}$).*

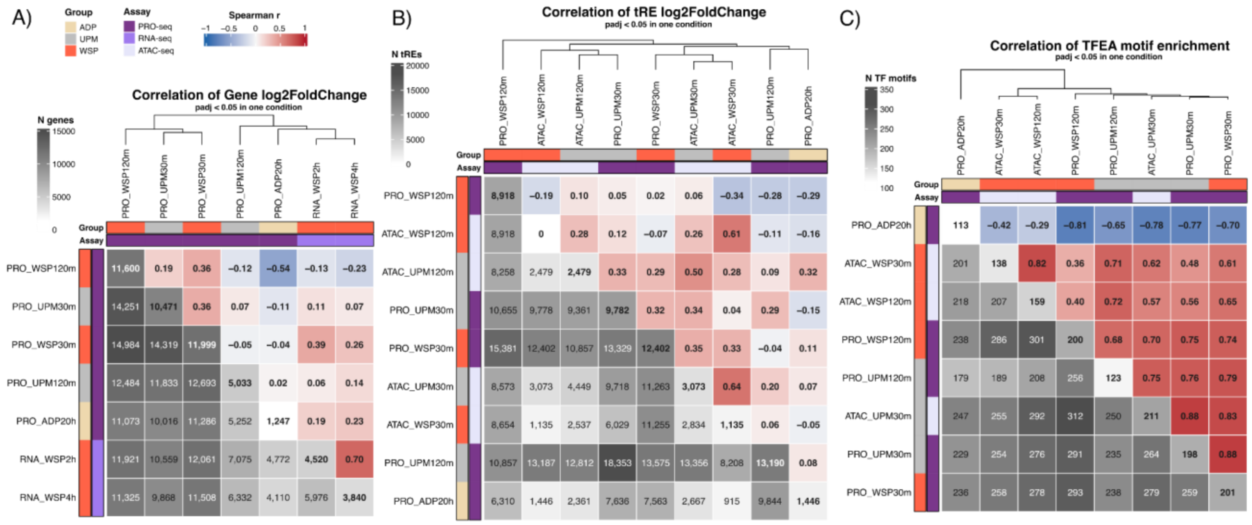


***Supplemental Figure Corr. Correlation of tRE, gene, and TF activity across assays.*** *A) Upper-right triangle shows spearman correlation of log2 fold change of gene expression (RNA-seq) and transcription (PRO-seq) across perturbations (WSP, UPM, ADP) and time-points. For each pairwise comparison, only tREs with DESeq2 adjusted-pvalue < 0.05 in at least one condition are considered (N features lower-left triangle). Diagonal shows N tREs with DESeq2 adjusted-pvalue < 0.05 in that condition. B) Same as A) but comparing tRE accessibility (ATAC-seq) and transcription (PRO-seq). C) Same as A) but comparing the GC-corrected enrichment scores (from TFEA-LE) of TF motifs, where GC-corrected adjusted p-value is used for the significant cut-off. All are compared to the appropriate vehicle. TFEA motif enrichments generally have higher correlation coefficients due to the lower number of features required to be compared. Results considering all features, regardless of significance, show comparable results.*

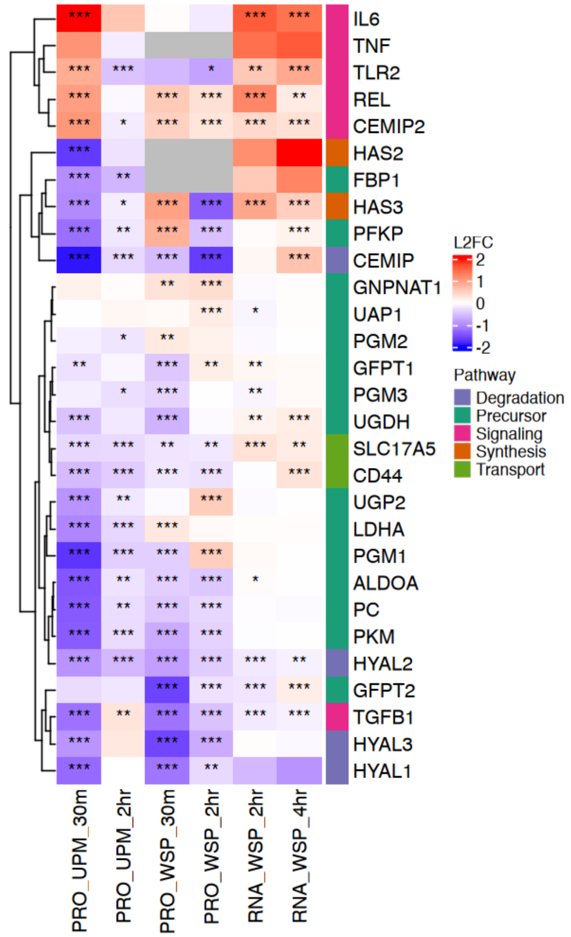


***Supplemental Figure HYA. Hyaluronan-related genes show similar regulation across WSP and UPM and steady-state.*** *Log2 fold change (Log2FC) compared to vehicle of smAEC cells perturbed with UPM (urban particulate matter) or Beas-2B cells perturbed with WSP (wood smoke particles) of genes involved in hyarluronan-related processes (Pathway). Adjusted p-values from DESeq2 are shown as * < 0.05, ** < 0.01, *** <0.001. PRO refers to PRO-seq, RNA refers to RNA-seq. Grey indicates that the gene was removed from analysis by DESeq2 due to low-expression outliers.*

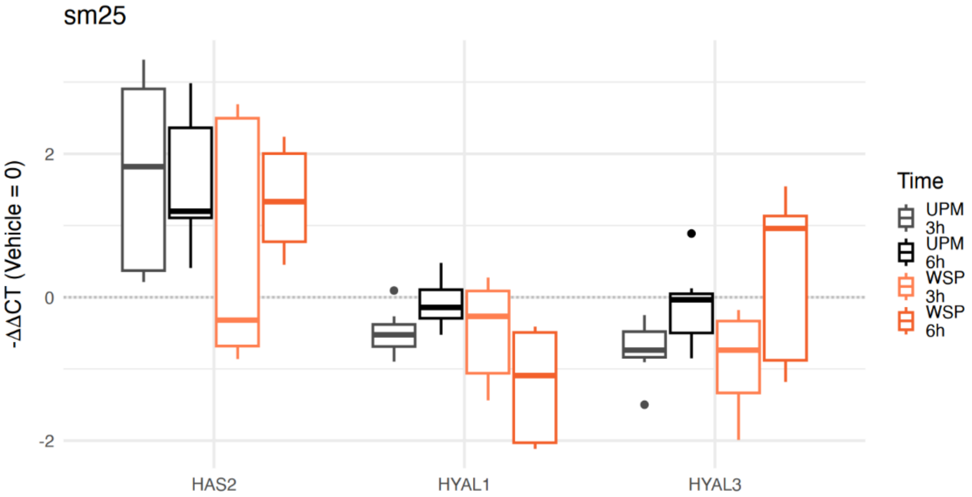

Supplemental Figure HYA_qRTPCR. **HYA synthesis genes generally go up upon pollutants while HYA degradation genes go down**. qRT-PCR results for smAECs treated with UPM and WSP, with results from RNA extracted at 3h or 6h time-points after perturbation compared to Vehicle. There are 8 replicates per condition/time-point, split between two different experiments/days.

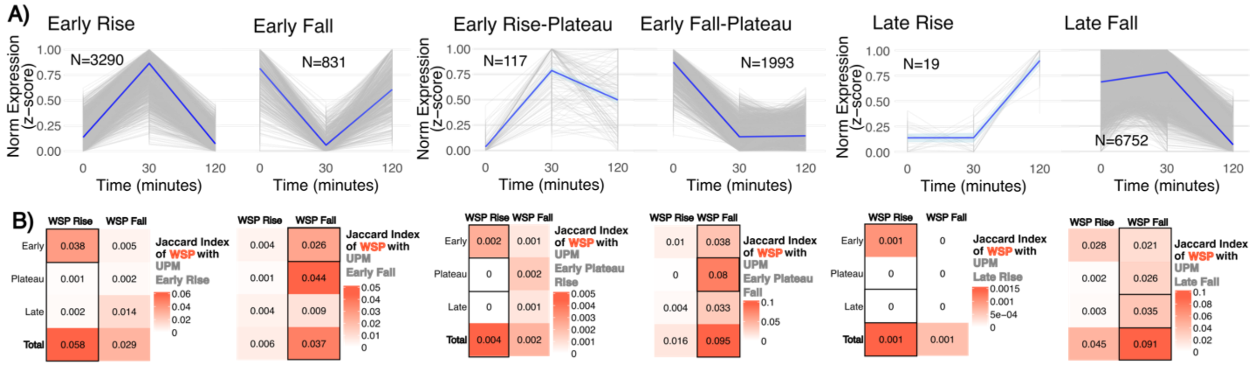


***Supplemental Figure EnhancerResponse. tREs show a large early response to UPM exposure comparable to genes and a unique large fall in expression at later points. A)*** *Line plots of the mean (blue, with light blue 95% confidence interval) and individual (grey) normalized transcriptional changes of tREs statistically significant (adjusted-p-value < 0.001) matching different timeline categories (details in Methods). Norm Expression (z-score) refers to normalized counts min-max scaled. 0 minutes refers to Vehicle response.* ***B).*** *Jaccard index for significant UPM tREs across the timeline categories of WSP responsive tREs (adjusted-p-value < 0.001).*

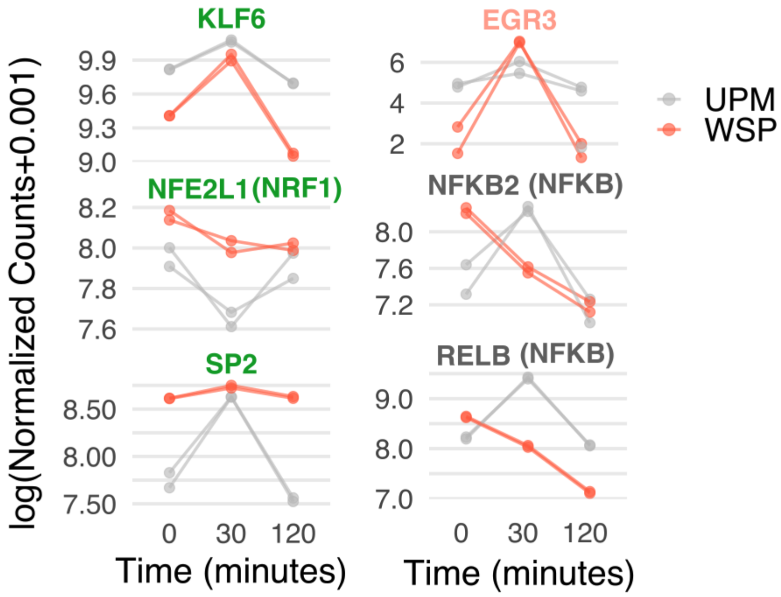


***Supplemental Figure TFlevels. Transcription factor transcriptional changes follow changes captured by motif analysis with TFEA.*** KLF6, NRF1, and SP2 all have motifs enriched in tREs with decreased transcription levels at the 30-minute mark according to TFEA in both WSP and UPM (green) and also show significant change in transcription for both perturbations at 30 minutes. KLF6 and SP2 have been observed both as activators and repressors. EGR3 only had significant negative enrichment score in WSP and shows significant upregulation in WSP. Genes encoding for the NFKB TFs show upregulation at 30 minutes only in UPM and have positive motif enrichment in UPM.

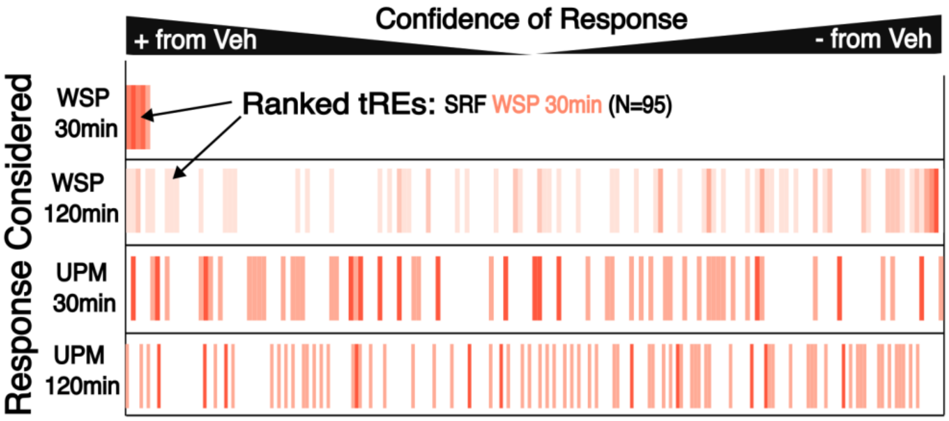


***Supplemental Figure SRFtREs. tREs attributed to SRF response in WSP response show no pattern of response in UPM.*** tREs called responding via SRF to WSP at 30 minutes are colored, and the x axes represent where all tREs rank in confidence of up(+) or down(-) regulation compared to Vehicle across 4 different responses: WSP 30min, WSP 120min, UPM 30min, UPM 120min. Left (or Right) most x-coordinates are tREs with the highest positive (or negative) log2fold change and lowest p-values for the listed response.

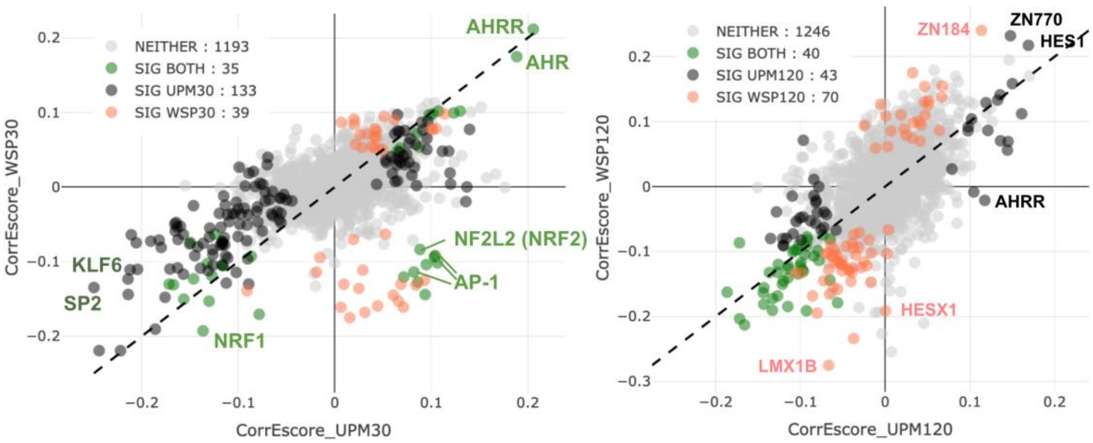


***Supplemental Figure TFEAscatter.*** **Timing dynamics of TF responses differ between WSP and UPM based on TFEA GC-corrected enrichment scores.** Transcription factors are highlighted as being a call in neither UPM/WSP, only one or the other, or both (where significance requires adjusted p-value < 0.01 and Fraction above Background < 0.46 – see Methods). **A)** 30 minutes compared to vehicle for ATAC-seq in nasal airway epithelial cells. Both KLF6 and SP2 have fraction above backgrounds of 0.51 in WSP, indicating weaker but still significant enrichment. **B)** WSP and UPM 120 minutes compared to vehicle for PRO-seq in small airway epithelial cells (UPM) or Beas-2B cells (WSP).

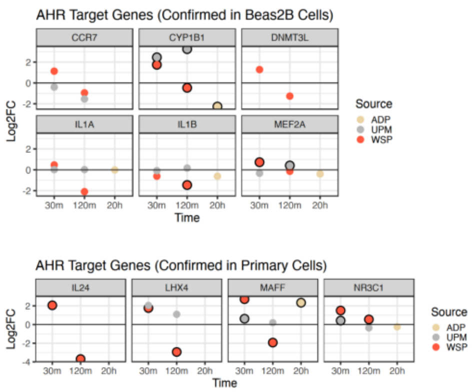


***Supplemental Figure AhRTargets. Previously confirmed AhR target genes show mostly similar responses at 30 minutes for UPM and WSP.*** *Log2 fold change (Log2FC) compared to vehicle of cells perturbed with ADP (Afghan dust particles), UPM (urban particulate matter), or WSP (wood smoke particles) of genes that were confirmed as AhR target genes based on ChIP-seq and sRNA knockdown of AhR in either Beas2B cells or small airway epithelial cells in Gupta et al. 2021. Dots with black outline indicate the gene has an adjusted p-value for fold change lower than* $1x10^{-10}$ *for WSP and UPM and 0.01 for ADP.*

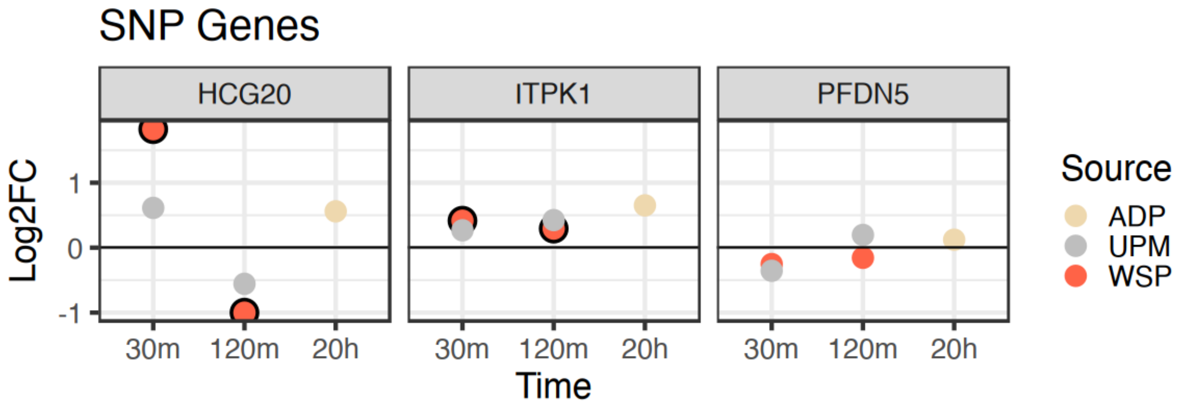


***Supplemental Figure SNP Genes. Three genes in which significantly associated SNPs are found tend to have changing transcriptional patterns shared across particulate perturbations****.* *DESeq-calculated log2 fold change (Log2FC) of genes within samples perturbed with one of three particulate matters (ADP=Afghan dust particles, UPM=urban particulate matter, WSP=wood smoke particles) at three time points (30 or 120 minutes, 20 hours) compared to vehicle sample. Black outline surrounds genes that have an adjusted p-value lower than 0.01.* PFDN5 has adjusted p-values all above 0.1. ITPK has adjusted p-values below 0.001 for UPM.

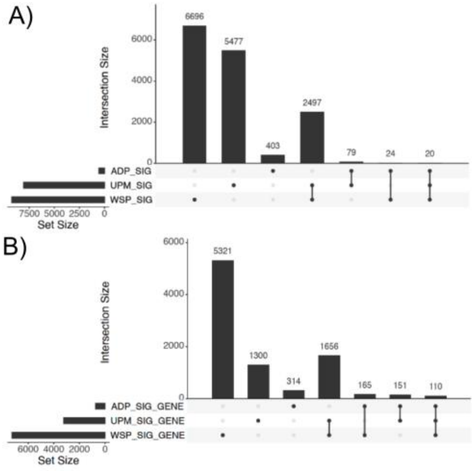


***Supplemental Figure UpSetResponse. Most tREs and genes responding according to strict significance cutoffs are unique to perturbations.*** *UpSet plots of A) tREs and B) genes called significantly responding at any time point for each perturbation compared to vehicle: ADP (Afghan dust particles 20h), UPM (urban particulate matter 30min or 120min), WSP (wood smoke particles 30min or 120min). Adjusted p-value cutoffs of 0.001 and 0.01 were used for tREs for UPM (N=8073) and WSP (N=9237), and ADP (N=526), respectively. Adjusted p-value cutoffs of $1x10^{-10}$ and 0.01 were used for genes for UPM and WSP, and ADP, respectively. Details are found in Methods.*

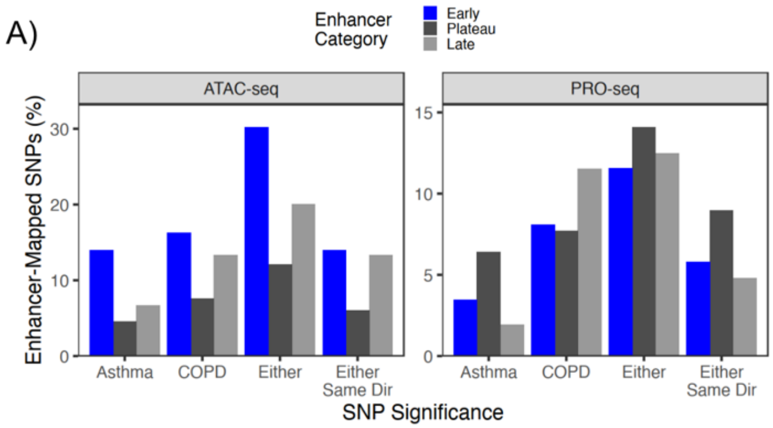


***Supplemental Figure SNP_sig****. Percentage of SNPs in enhancer timing categories (agreed upon between WSP and UPM) based on ATAC-seq and PRO-seq that are associated with asthma or COPD.*

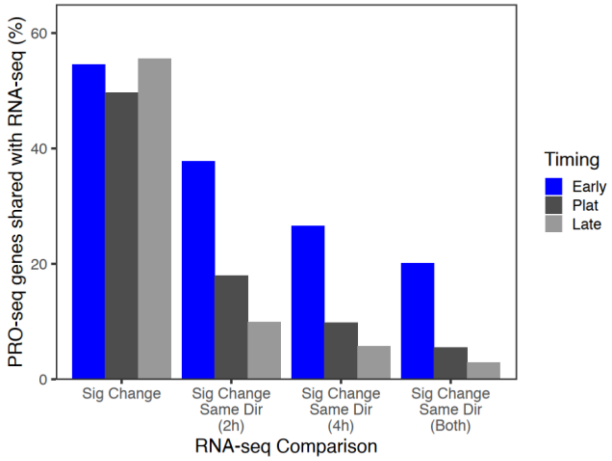


*Supplemental Figure RNAseq_Comp.* ***Harmonization between RNA-seq and PRO-seq responsive genes is not biased against early response genes.*** *Percentage of genes responding in the three time categories in PRO-seq (early fall/rise (Early), early plateau fall/rise (Plat), late fall/rise (Late) that have a significant change in RNA-seq (Sig Change) in the same direction at the 2h, 4h, or both timepoints. Beas-2B cells perturbed with WSP were used for both.*

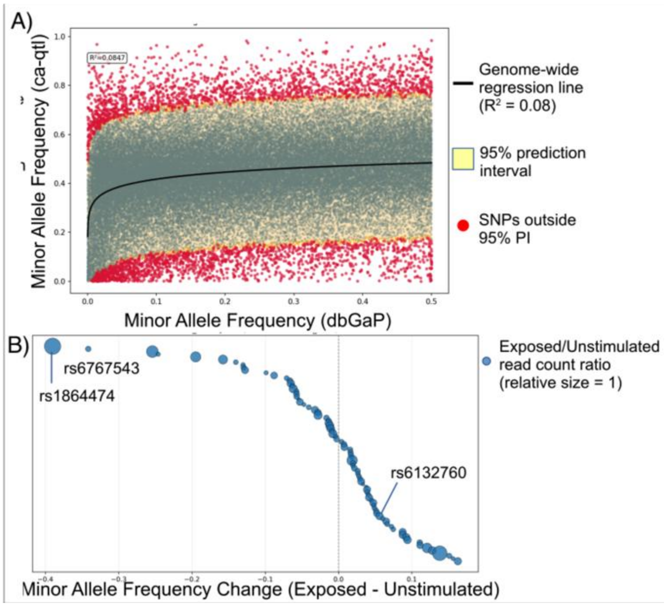


***Supplemental Figure CAQTL****. A) Genome-wide distribution of SNPs based on counted minor allele frequency vs expected minor allele frequency in dbGaP. Allele-specific reads were counted at 68,204 SNPs in 262,981 consensus ATAC-seq peaks and the minor allele frequency was compared with the population minor allele frequency in dbGaP. The black line represents a 4-parameter logistic regression for the data and the yellow shading represents a 95% prediction interval calculated around the regression line. Each red dot represents a SNP that falls outside the prediction interval. Blue dots represent SNPs inside the prediction interval.  B) Bubble plot showing the change in minor allele frequency between exposed and unstimulated ca-qtl analyses. Negative values represent a decrease in minor allele frequency with exposed samples. Bubble size is a relative representation of normalized read count change between exposed and*

*unstimulated samples as a fraction (exposed/unstimulated). The legend contains a reference bubble with a value of 1.*

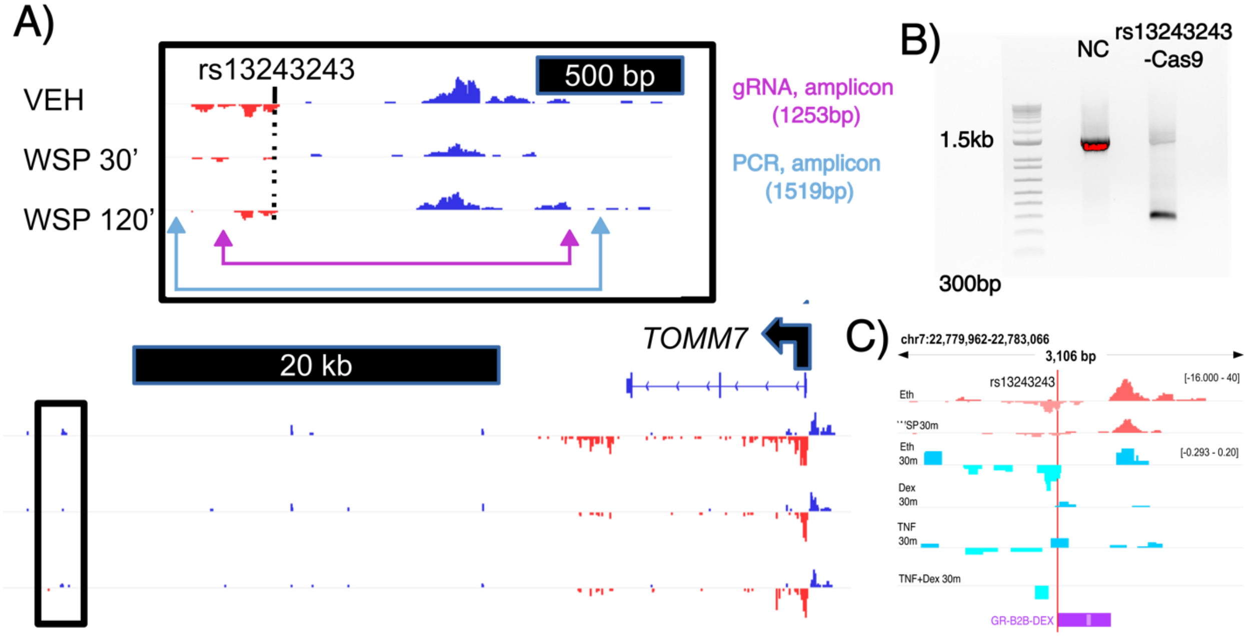


***Supplemental Figure TOMM7SNP**. A) Location/length of gRNA/PCR primers for Cas9-editing and location of enhancer compared to TOMM7 based on IGV tracks of PRO-seq of Beas-2B cells perturbed with wood smoke particles (WSP). B. Gel from PCR amplification for negative control (NC) and the knockdown genotype. C) IGV tracks of PRO/GRO-seq of Beas-2B cells perturbed with WSP, TNF, and/or dexamethasone (Dex) (Cond column); the numerical read distributions are not directly comparable outside of experiments due to different normalization approaches (scale noted on the right top of each experiment). The SNP location and ChIP peaks from Beas2B cells perturbed with Dex (GR-B2B-DEX) are highlighted.*

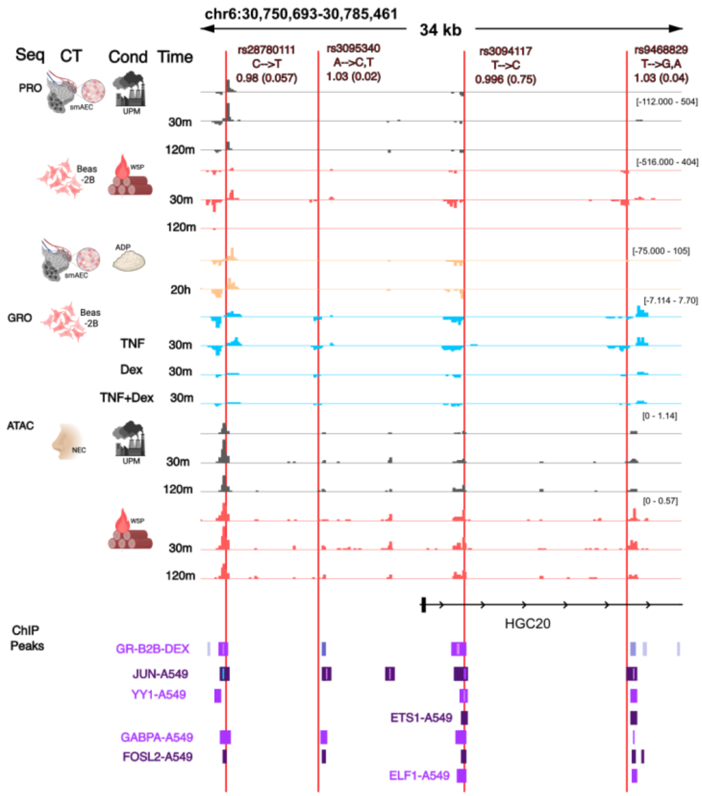


***Supplemental Figure Dex SNP. Previous SNP correlated with asthma is within group of tREs and other SNPs similarly associated.*** *IGV tracks of PRO/GRO-seq or ATAC-seq (Seq column) of lung cells (small airway epithelial cells (smAEC), Beas-2B cells, nasal epithelial cells (NEC)) (CT column) perturbed with urban particulate matter (UPM), wood smoke particles (WSP), afghan dust particles (ADP), TNF, and/or dexamethasone (Dex) (Cond column) at varying time points (Time column). The numerical read distributions are not directly comparable outside of experiments due to different normalization approaches (scale noted on the right top of each experiment). ChIP peaks from A549 (ENCODE) of different transcription factors or from Beas2B cells perturbed with Dex (GR-B2B-DEX) are highlighted with varying colors for easier interpretation. The following ENCODE files were used: JUN=ENCFF618CNL, YY1=ENCFF835PPK, ETS1=ENCFF271WHK, GABPA=ENCFF343BIK, FOSL2=ENCFF232TVP, ELF1=ENCFF169QCM. SNPs are labeled by their rsid values and odds ratios for asthma (p-values) in our study.*

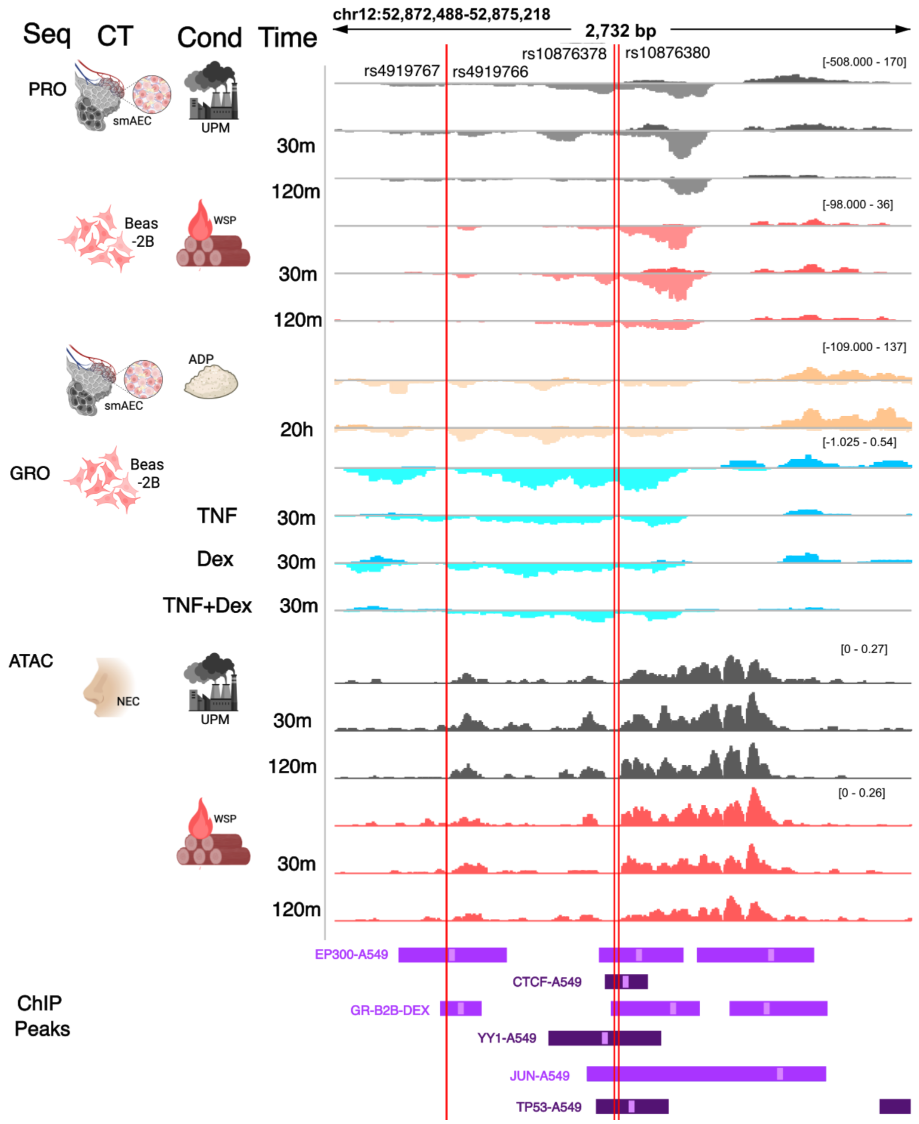


*Supplemental Fig Chr12SNPs.* ***Chromosome 12 SNPs fall within enhancer group with consistent signal across conditions and omics data****. IGV tracks of PRO/GRO-seq or ATAC-seq (Seq column) of lung cells (small airway epithelial cells (smAEC), Beas-2B cells, nasal epithelial cells (NEC)) (CT column) perturbed with urban particulate matter (UPM), wood smoke particles (WSP), afghan dust particles (ADP), TNF, and/or dexamethasone (Dex) (Cond column) at varying time points (Time column). The numerical read distributions are not directly comparable outside of experiments due to different normalization approaces (scale noted on the right top of each experiment). ChIP peaks from A549 (ENCODE) of different transcription factors or from Beas2B cells perturbed with Dex (GR-B2B-DEX). The following ENCODE files were used: EP300=ENCFF096PTH, CTCF=ENCFF229ULU, YY1=ENCFF835PPK, JUN=ENCFF618CNL, TP53=ENCFF229ULU. SNPs are labeled by their rsids*

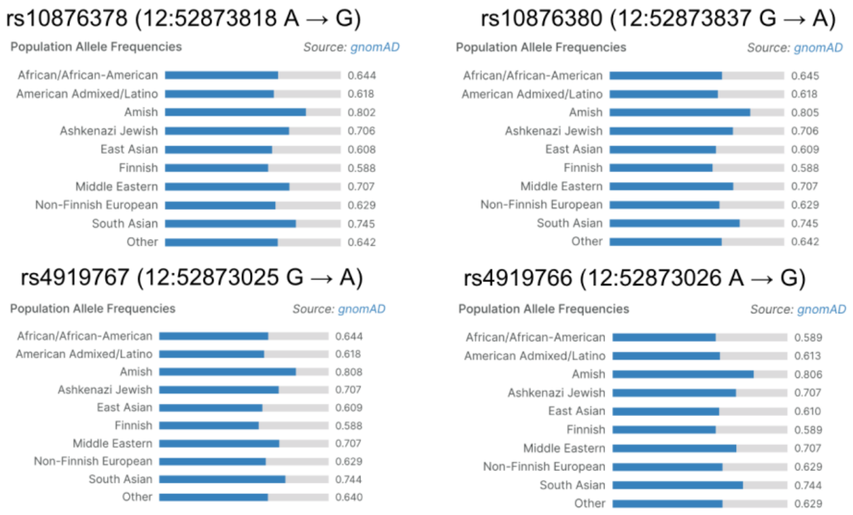


***Supplemental Figure Ppln. Four of the top SNPs indicating lower likelihood of COPD and asthma are more common across all ancestry populations and at similar levels.*** *Population allele frequencies from Open Targets webserver (https://platform.opentargets.org/) of each of the four SNPs falling within chromosome 12 and within the top 10 SNPs associated with both COPD and asthma.*
